# Factors Associated with COVID-19 Breakthrough Infection in the Pre-Omicron Era Among Vaccinated Patients with Rheumatic Diseases: A Cohort Study

**DOI:** 10.1101/2022.07.13.22277606

**Authors:** Naomi J. Patel, Xiaosong Wang, Xiaoqing Fu, Yumeko Kawano, Claire Cook, Kathleen M.M. Vanni, Grace Qian, Emily Banasiak, Emily Kowalski, Yuqing Zhang, Jeffrey A. Sparks, Zachary S. Wallace

## Abstract

**Objective:** Rheumatic disease patients on certain immunomodulators are at increased risk of impaired humoral response to SARS-CoV-2 vaccines. We aimed to identify factors associated with breakthrough infection among patients with rheumatic diseases.

**Methods:** We identified patients with rheumatic diseases being treated with immunomodulators in a large healthcare system who received at least two doses of either the mRNA-1273 (Moderna) or BNT162b2 (Pfizer-BioNTech) vaccines or one dose of the Johnson & Johnson-Janssen (J&J) vaccine. We followed patients until SARS-CoV-2 infection, death, or December 15, 2021, when the Omicron variant became dominant in our region. We estimated the association of baseline characteristics with the risk of breakthrough infection using multivariable Cox regression.

**Results:** We analyzed 11,468 patients (75% female, mean age 60 years). Compared to antimalarial monotherapy, multiple immunomodulators were associated with higher risk of infection: anti-CD20 monoclonal antibodies (aHR 5.20, 95% CI: 2.85, 9.48), CTLA-4 Ig (aHR 3.52, 95% CI: 1.90, 6.51), mycophenolate (aHR 2.31, 95% CI: 1.25, 4.27), IL-6 inhibitors (aHR 2.15, 95% CI: 1.09, 4.24), JAK inhibitors (aHR 2.02, 95% CI: 1.01, 4.06), and TNF inhibitors (aHR 1.70, 95% CI: 1.09, 2.66). mRNA-1273 recipients had a lower risk of breakthrough infection compared to BNT162b2 recipients (aHR 0.66, 95% CI: 0.50, 0.86). There was no association of sex, body mass index, smoking status, race, or ethnicity with risk of breakthrough infection.

**Conclusion:** Among patients with rheumatic diseases, multiple immunomodulators were associated with increased risk of breakthrough infection. These results highlight the need for additional mitigation strategies in this vulnerable population.

## Introduction

SARS-CoV-2 vaccines are highly effective against severe outcomes even among immunosuppressed patients, but breakthrough infections after vaccination have been observed.^1-4^ There is particular concern among patients with systemic autoimmune rheumatic diseases (SARDs) that some may be at increased risk of breakthrough infection because of underlying immune dysregulation, immunomodulatory or immunosuppressive medications, and comorbid conditions, which may contribute to decreased vaccine efficacy and predisposition to breakthrough infection.^5-9^ Indeed, a number of medications commonly used to treat immune-mediated diseases, including anti-CD20 monoclonal antibodies, antimetabolites (e.g., methotrexate, mycophenolate), glucocorticoids, and cytokine inhibitors (e.g., TNF inhibitors), have been associated with impaired humoral response to SARS-CoV-2 and other vaccines.^10-14^

While a number of studies have evaluated the immunologic response to vaccination, there are limited data on the risk of breakthrough infection in patients with SARDs. Case series in the early vaccine era described breakthrough infections, including severe disease, especially in patients receiving B cell depletion, mycophenolate, and methotrexate.^7,15,16^ More recently, there have been conflicting reports regarding the association of SARDs and their treatments with the risk of breakthrough infection.^17-19^ Based on these evolving data, the United States Centers for Disease Control and Prevention continues to revise their vaccine recommendations for immunocompromised patients who are now advised to receive additional vaccine doses.^20^

To guide management of this vulnerable population during the ongoing pandemic, additional data regarding risks for COVID-19 breakthrough infections among patients with SARDs are needed. We aimed to evaluate the baseline factors associated with COVID-19 breakthrough infections in patients with SARDs in a large healthcare system in the United States.

## Methods

### Study population and design

We performed a retrospective cohort study investigating factors associated with breakthrough infection among patients with SARDs at a large healthcare system. Mass General Brigham (MGB) is a multi-center healthcare system that includes a total of 14 hospitals, including two tertiary care hospitals (Massachusetts General Hospital and Brigham and Women’s Hospital), as well as other primary care and specialty outpatient centers in the greater Boston, Massachusetts area. We identified SARS-CoV-2 vaccine recipients from within MGB who were ≥18 years of age, Massachusetts residents, and had a SARD diagnosis. This study was approved by the MGB Institutional Review Board (2020P000833).

### Identification of vaccinated patients and index date

We limited our study population to Massachusetts residents (as obtained by primary address in the electronic health record [EHR]) because vaccination data from the Massachusetts state database was populated in the EHR. All immunizations administered in Massachusetts are required by law to be reported to the Massachusetts Immunization Information System.^21^ We identified all patients in the MGB EHR who were fully vaccinated according to the earliest definitions used by the CDC: two doses of a messenger ribonucleic acid (mRNA) SARS-CoV-2 vaccine (i.e., either BNT162b2 [Pfizer-BioNTech] or mRNA-1273 [Moderna]) or one dose of the Ad26.COV2.S (Johnson & Johnson-Janssen) vaccine, between December 11, 2020 (when the first vaccine was initially approved under emergency use authorization in the US) and November 15, 2021. This definition continues to inform the definition of “fully vaccinated” used by health agencies, among others. Although immunosuppressed patients are eligible for an additional dose to complete their primary vaccine series, the majority of people in the US have only received this initial vaccine series to date, despite recommended additional doses and boosters.^22^ We excluded patients who received only one dose of an mRNA vaccine, those who received one dose of an mRNA vaccine followed by a dose of the Johnson & Johnson-Janssen vaccine, and those whose vaccine type was unknown. We considered the index date to be the date of the second dose of either mRNA vaccine or the date of the single Johnson & Johnson-Janssen vaccine.

We also identified additional vaccine doses received after the index date. However, these were not considered as baseline factors in this analysis since they occurred after the index date.

### Identification of systemic autoimmune rheumatic diseases

From this cohort of vaccinated individuals, we identified patients with SARDs (**Figure 1**). To do so, we required that patients have: 1) Two International Classification of Diseases (ICD)-10 codes (**Supplementary Table 1**) for a SARD diagnosis within two years prior to the index date, one of which needed to be in the one year prior to the index date, and separated by at least 30 days, and 2) receipt preceding the index date of an immunomodulator, which we defined as either a prescription for a conventional synthetic, targeted, or biologic disease modifying anti-rheumatic drug (DMARD) (**Supplementary Table 2**) within 12 months of the index date or a prescription for ≥30 pills of either oral prednisone or methylprednisolone within 6 months of the index date. We required all patients to have received immunomodulatory treatment to enhance the specificity of the algorithm and to examine the impact specific medications may have on risk for breakthrough infection. The positive predictive value (PPV) for a similar rules-based algorithm to identify rheumatoid arthritis was 86%.^23^ We reviewed 50 random patients who met our algorithm and 45 had physician-confirmed SARDs, confirming a PPV of approximately 90%. Patients with osteoarthritis, fibromyalgia, or crystalline arthritis without another concomitant SARD diagnosis were excluded as these are generally not considered to be SARDs and are not typically treated with long-term immunomodulators.

**Figure 1.**
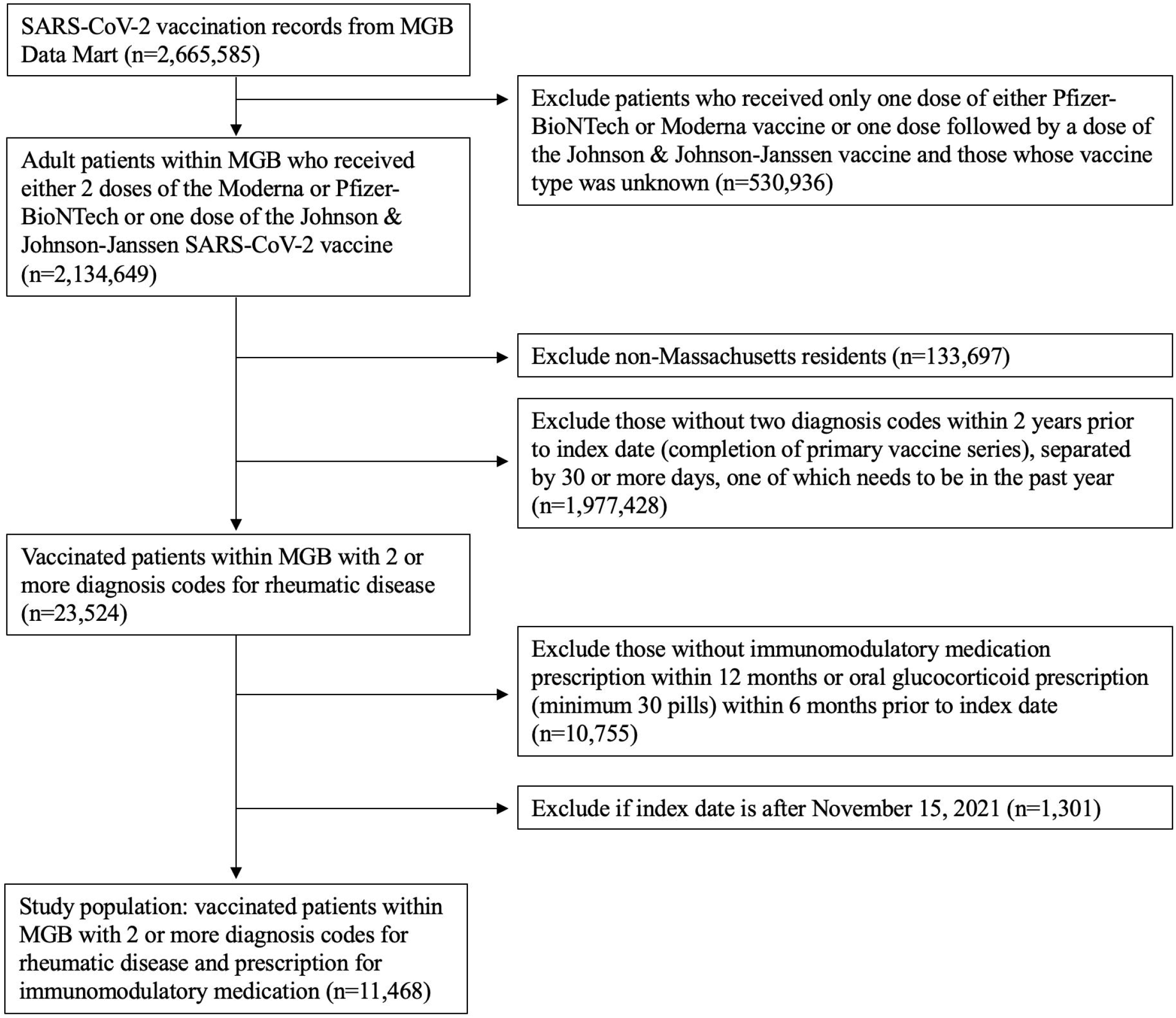
Identification of vaccinated rheumatic disease patients within the Mass General Brigham healthcare system. MGB, Mass General Brigham healthcare system

### Ascertainment of potential risk factors and covariates

Data regarding dates and presence of ICD codes, medication prescriptions, demographics, and comorbidities were extracted from the MGB electronic data warehouse, as previously described.^24^ The patient’s primary rheumatic disease diagnosis was based on ICD-10 codes. In some cases, patients had codes associated with multiple rheumatic diseases. In scenarios where one disease is often secondary to or associated with a primary condition (e.g., antiphospholipid syndrome in systemic lupus erythematosus), the patient was considered to have the primary disease (e.g., systemic lupus erythematosus). In cases where there was a discrepancy (e.g., giant cell arteritis and ANCA-associated vasculitis), the disease associated with the ICD code used most frequently was considered to be the primary diagnosis. Cases in which ICD codes that can coexist (e.g., seronegative spondyloarthropathy and giant cell arteritis) or can exist as overlap disease (e.g., systemic lupus erythematosus and rheumatoid arthritis) were categorized as “multiple” primary rheumatic diseases. Since many patients had ICD codes for both giant cell arteritis and polymyalgia rheumatica, we considered this as a single combined category.

Baseline characteristics including demographics (including race/ethnicity as obtained from the electronic health record), comorbidities as defined by ICD-10 codes, smoking history, and body mass index (BMI) were extracted from the electronic data warehouse and assessed in the one year prior to the index date. The Charlson Comorbidity Index (CCI) was calculated using all available data from comorbidities as ascertained by ICD-10 in the one year prior to the index date.^25^ We also assessed the type of vaccine received as part of the initial vaccine series used in our case definition (Moderna, Pfizer-BioNTech, or Johnson & Johnson-Janssen). Last, we assessed receipt of an additional dose of the SARS-CoV-2 vaccine, defined as the second of any vaccine for those who received the Johnson & Johnson-Janssen vaccine initially or the third of any vaccine for those who received two doses of either the Pfizer-BioNTech or Moderna vaccines.

### Outcome assessment: breakthrough infection

The primary outcome was SARS-CoV-2 breakthrough infection, defined as: 1) a positive SARS-CoV-2 polymerase chain reaction (PCR) or antigen test from a nasopharyngeal or respiratory specimen, and/or 2) a positive COVID-19 flag in the EHR on or after the index date. In MGB, a COVID-19 flag indicates a confirmed diagnosis of SARS-CoV-2 infection and captures patients with a confirmed positive test outside of our healthcare system. We also included patients flagged as having COVID-19 based on a positive home rapid antigen assay reported to providers or clinics. In some cases, results from tests performed outside of MGB were automatically pulled into the electronic data warehouse because of a linkage across other healthcare systems. Outcomes in the primary analysis were assessed from the index date through December 15, 2021, when the Omicron variant became the dominant strain in Massachusetts.^26^ A sensitivity analysis extended follow-up through February 22, 2022, including initial phases of the Omicron wave. As a secondary outcome, we also identified patients who experienced severe outcomes defined as hospitalization or death within 30 days of the initial positive test.

### Statistical analysis

Categorical variables are presented as number (percentage), and continuous variables are presented as mean ± standard deviation or median ± interquartile range, as appropriate. The index date was defined as the date of the second dose of an mRNA SARS-CoV-2 vaccine (either Moderna or Pfizer-BioNTech) or the first dose of the Johnson & Johnson-Janssen vaccine. Patients were followed from the index date until the earliest date of: 1) death, 2) the first positive SARS-CoV-2 PCR, antigen test, or COVID-19 flag in the EHR, or 3) December 15, 2021, and person-months of follow-up were determined for each patient.

We assessed the association of factors of interest among SARD patients with the risk of breakthrough infection using unadjusted and multivariable Cox regression models to estimate hazard ratios (HR) and 95% confidence intervals (CIs). For the purpose of categorizing patients into a single medication group in the Cox regression models, antimalarial monotherapy was used as the reference group. For those on multiple medications, we considered a hierarchical system to categorize each patient into a single category as follows: anti-CD20 monoclonal antibody, cyclophosphamide, tumor necrosis factor (TNF) inhibitor, IL-6 inhibitor, cytotoxic T-lymphocyte associated protein 4 immunoglobulin (CTLA-4 Ig), IL-17 inhibitor/IL-23 inhibitor/IL-12/23 inhibitor, other bDMARD, tsDMARD/Janus kinase (JAK) inhibitor, mycophenolate mofetil or mycophenolic acid, methotrexate, then other csDMARD. For example, a patient receiving both a TNF inhibitor and methotrexate would be classified as being on a TNF inhibitor for the purpose of our primary analysis. The multivariable model was adjusted for age, sex, smoking status, CCI, and vaccine type based on findings from univariate analyses and directed acyclic graphs of known risk factors for breakthrough infection.

To assess the robustness of our results, we performed several subgroup and sensitivity analyses in which we: 1) Only considered breakthrough infection outcomes as those occurring at least 14 days after the index date (thus, those who would be considered to have breakthrough infections based on the definitions by the Centers for Disease Control & Prevention); 2) Censored at the time of any additional vaccine dose, defined as the second of any vaccine for those who received initial Johnson & Johnson-Janssen vaccine or third of any vaccine for those who received two of either the Pfizer-BioNTech or Moderna vaccines initially; and 3) Extended follow-up through February 22, 2022, thus including the initial period during which the Omicron variants became the local dominant strains.^27^

We assessed for proportional hazards assumption by including an interaction term between the exposure of interest and follow-up time. The proportional hazards assumption was met in all analyses. The level of significance was set as a two-tailed p<0.05, and statistical analyses were completed using SAS statistical software (version 9.4; SAS Institute, Inc., Cary, NC).

## Results

### Study sample

Among 2.1 million vaccinated patients within MGB, we identified 11,468 vaccinated patients (at least two mRNA vaccines or one Johnson & Johnson-Janssen vaccine) with SARDs prescribed immunomodulatory medications (**Figure 1**). The majority (8620, 75%) were female, and the mean age was 60 ± 15 years (**Table 1**). Most patients were White (9490, 83%), followed by Black (628, 6%), and Asian (417, 4%).

**Table 1.**
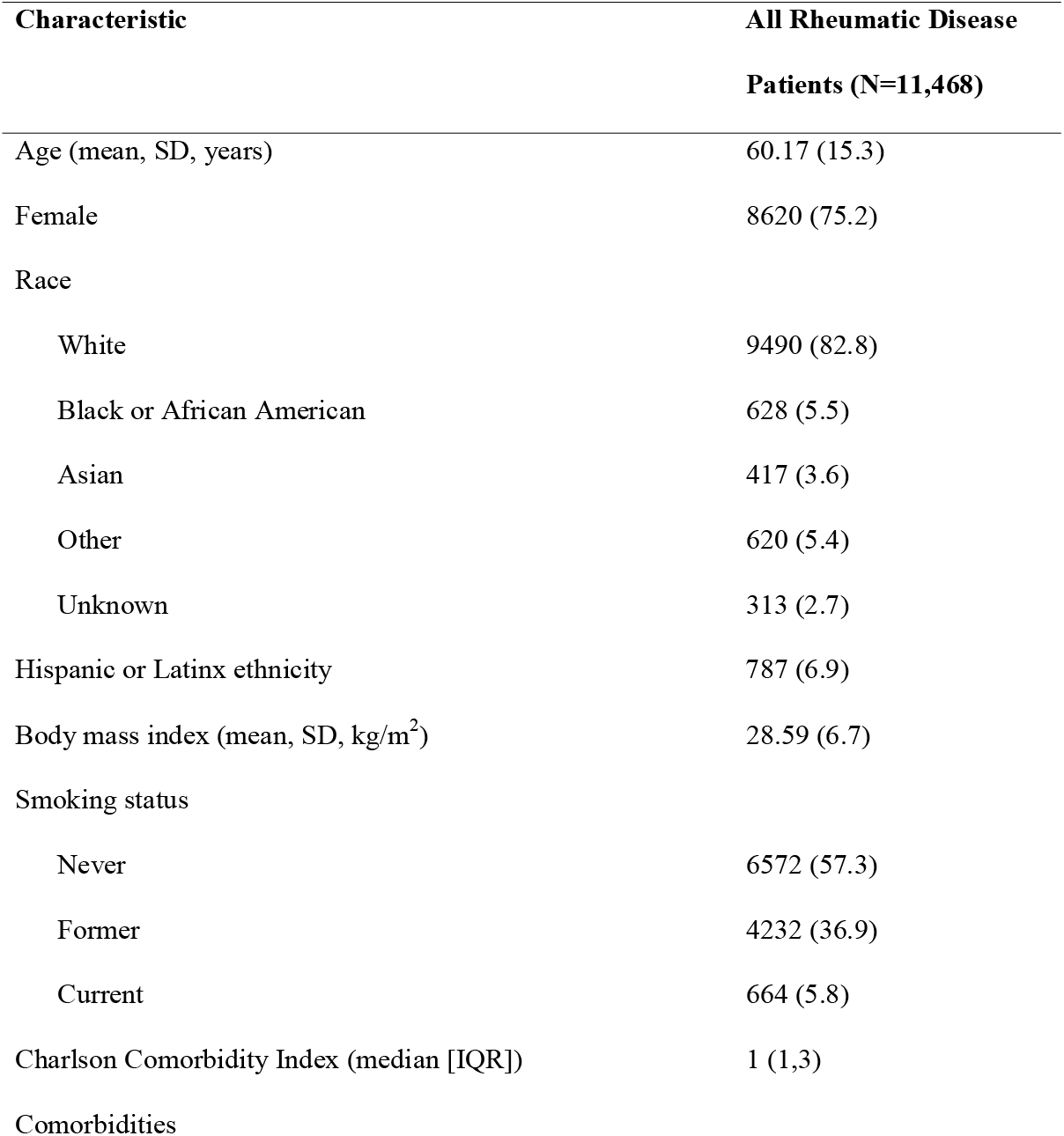
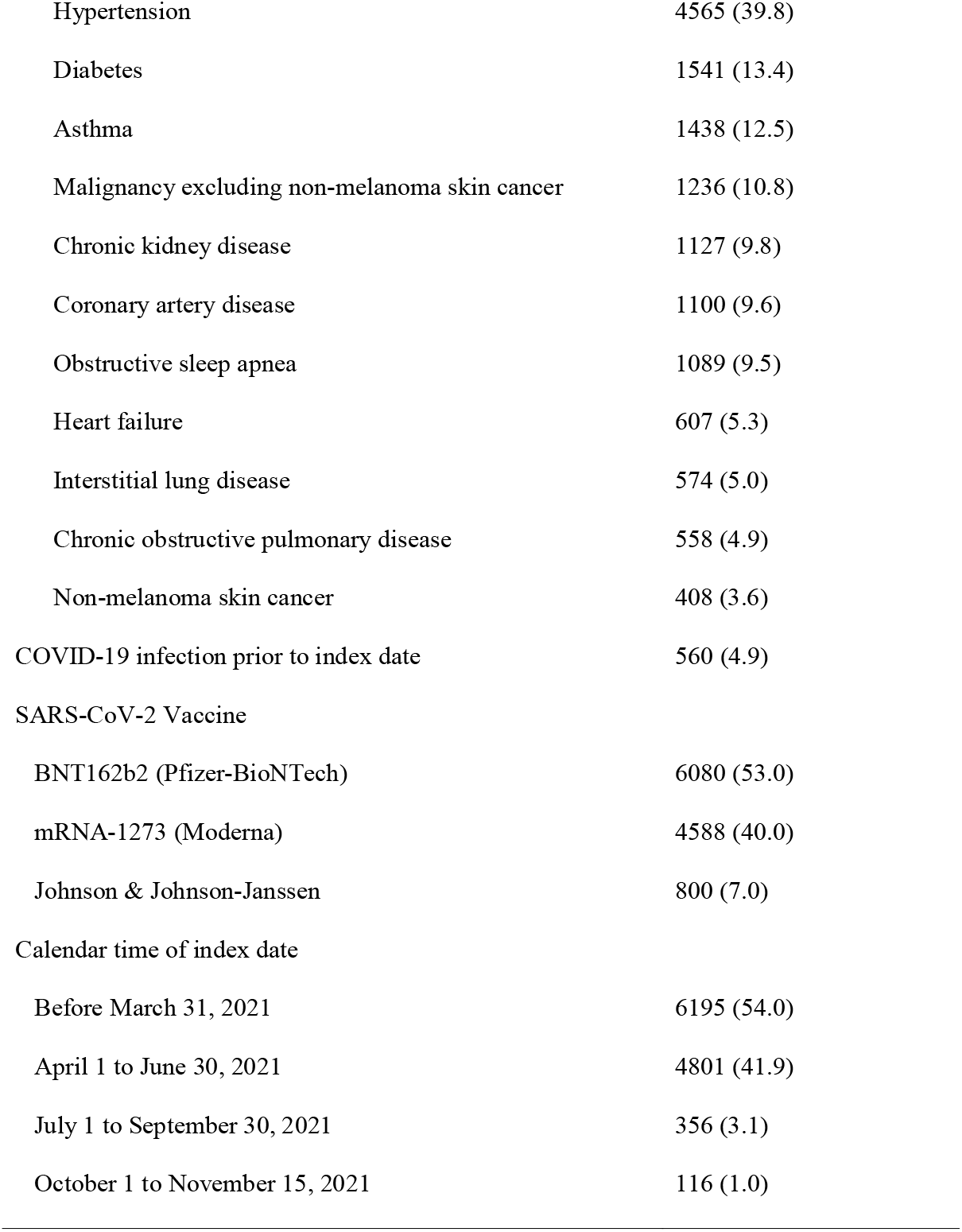
Demographic and clinical characteristics of patients with systemic rheumatic disease at the time of SARS-CoV-2 vaccination.

| Characteristic | All Rheumatic Disease Patients (N=11,468) |
| --- | --- |
| Age (mean, SD, years) | 60.17 (15.3) |
| Female | 8620 (75.2) |
| Race |  |
| White | 9490 (82.8) |
| Black or African American | 628 (5.5) |
| Asian | 417 (3.6) |
| Other | 620 (5.4) |
| Unknown | 313 (2.7) |
| Hispanic or Latinx ethnicity | 787 (6.9) |
| Body mass index (mean, SD, kg/m<sup>2</sup>) | 28.59 (6.7) |
| Smoking status |  |
| Never | 6572 (57.3) |
| Former | 4232 (36.9) |
| Current | 664 (5.8) |
| Charlson Comorbidity Index (median [IQR]) | 1 (1,3) |
| Comorbidities |  |
| Hypertension | 4565 (39.8) |
| Diabetes | 1541 (13.4) |
| Asthma | 1438 (12.5) |
| Malignancy excluding non-melanoma skin cancer | 1236 (10.8) |
| Chronic kidney disease | 1127 (9.8) |
| Coronary artery disease | 1100 (9.6) |
| Obstructive sleep apnea | 1089 (9.5) |
| Heart failure | 607 (5.3) |
| Interstitial lung disease | 574 (5.0) |
| Chronic obstructive pulmonary disease | 558 (4.9) |
| Non-melanoma skin cancer | 408 (3.6) |
| COVID-19 infection prior to index date | 560 (4.9) |
| SARS-CoV-2 Vaccine |  |
| BNT162b2 (Pfizer-BioNTech) | 6080 (53.0) |
| mRNA-1273 (Moderna) | 4588 (40.0) |
| Johnson & Johnson-Janssen | 800 (7.0) |
| Calendar time of index date |  |
| Before March 31, 2021 | 6195 (54.0) |
| April 1 to June 30, 2021 | 4801 (41.9) |
| July 1 to September 30, 2021 | 356 (3.1) |
| October 1 to November 15, 2021 | 116 (1.0) |

Common comorbidities included hypertension (4565, 40%), asthma (1438, 13%), diabetes (1541, 13%), coronary artery disease (1100, 10%), chronic kidney disease (1127, 10%), and obstructive sleep apnea (1089, 10%). Additionally, 1236 (11%) patients had a diagnosis of any malignancy excluding non-melanoma skin cancers. The median (IQR) Charlson Comorbidity Index was 1 (1, 3).

The most common SARDs were rheumatoid arthritis (6099, 53%), psoriatic arthritis (1777, 15%), systemic lupus erythematosus (1479, 13%), and giant cell arteritis and/or polymyalgia rheumatica (388, 3%) (**Table 2**). Among all patients, 3739 (33%) were on hydroxychloroquine, 4968 (43%) were on biologic DMARDs, 702 (6%) were on targeted synthetic DMARDs (i.e., JAK inhibitors), 8173 (71%) were on conventional synthetic DMARDs, and 1162 (10%) were on oral glucocorticoids. The most common biologic DMARDs included tumor necrosis factor inhibitors (3142, 27%) followed by IL-6 inhibitors (458, 4%) (**Table 2**). The most common conventional synthetic DMARDs aside from hydroxychloroquine included methotrexate (4058, 35%), followed by leflunomide (596, 5%) and sulfasalazine (536, 5%).

**Table 2.**
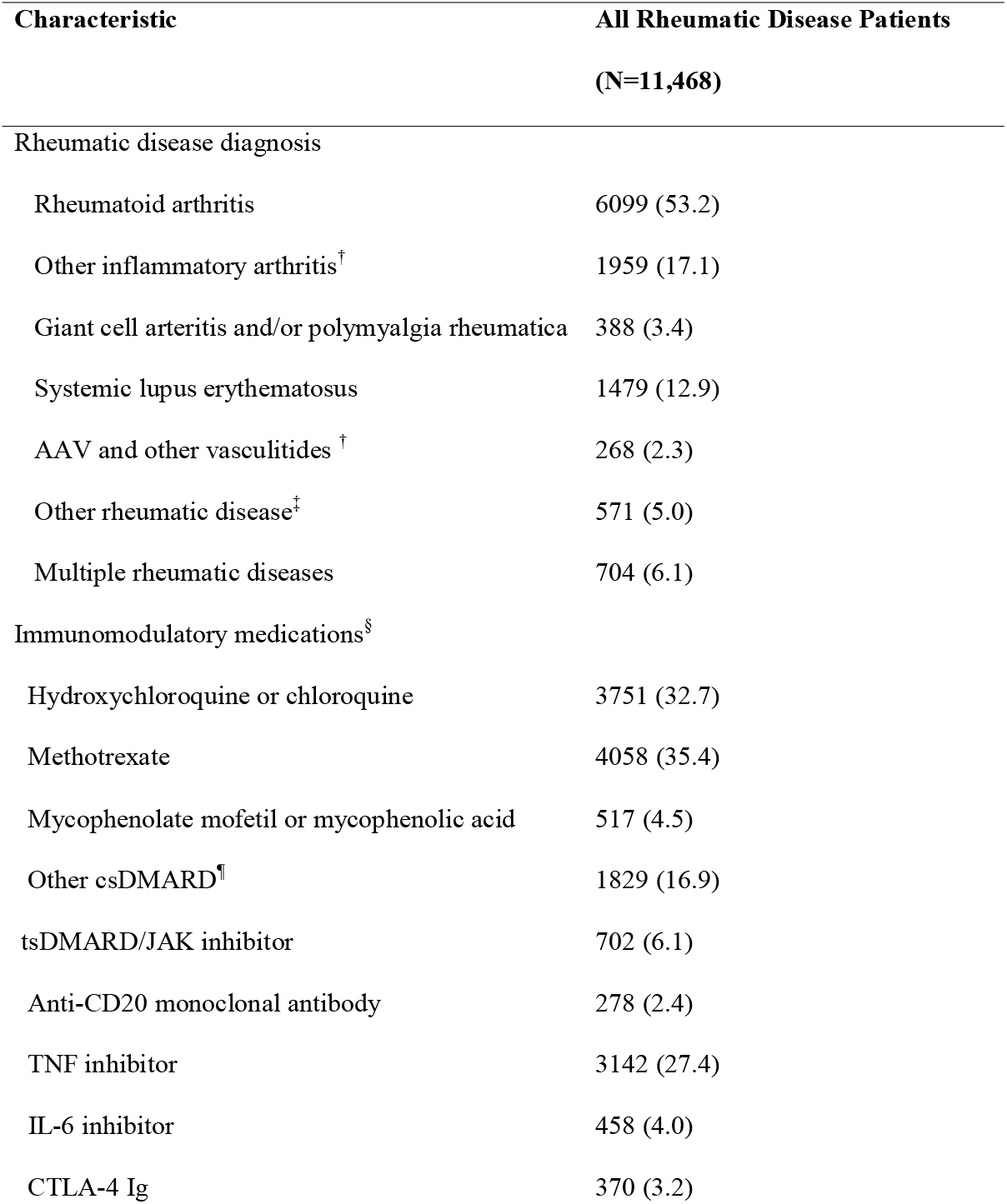
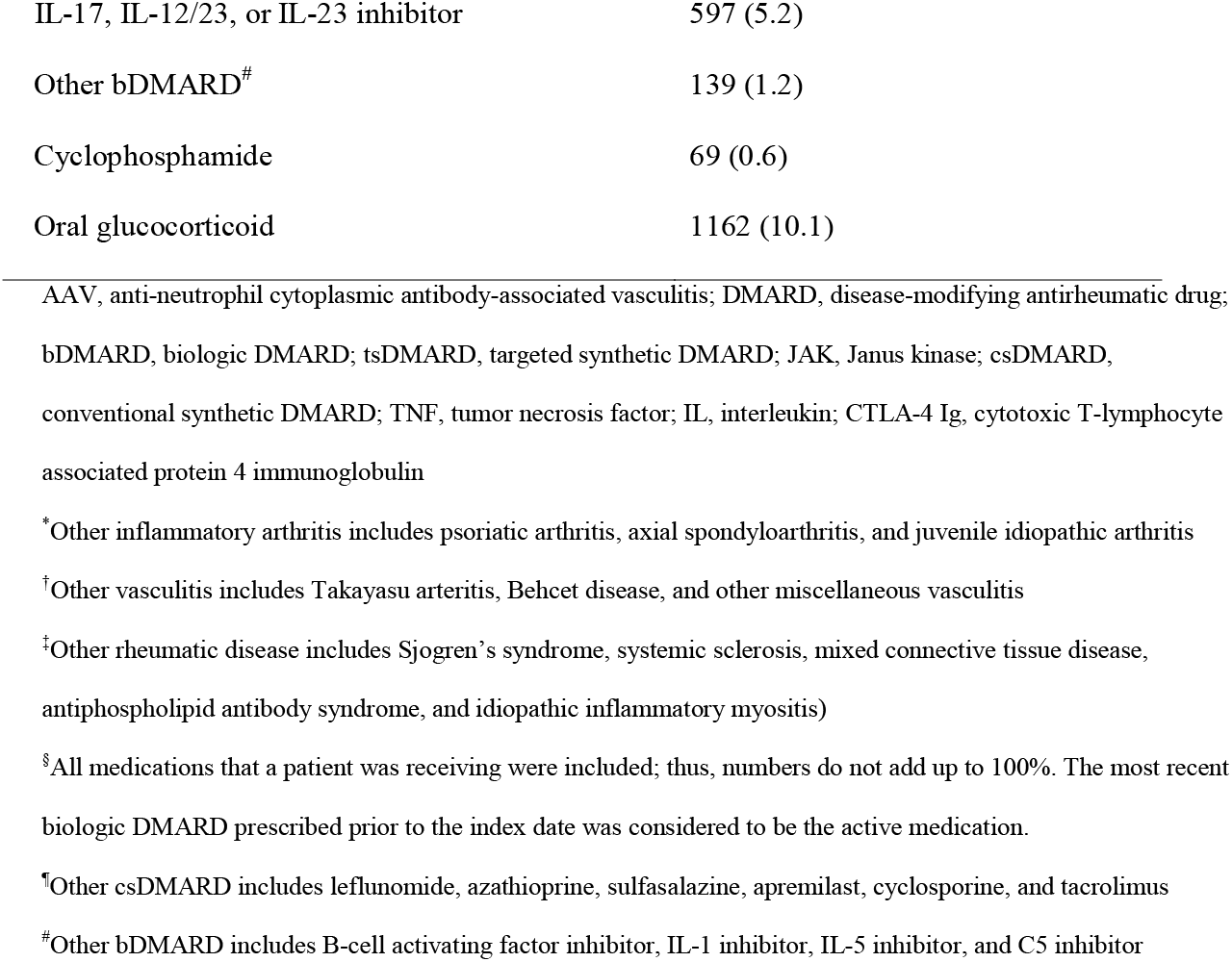
Rheumatic disease characteristics of patients at the time of SARS-CoV-2 vaccination.

| **Characteristic** | **All Rheumatic Disease Patients**<br>**(N=11,468)** |
| --- | --- |
| **Rheumatic disease diagnosis** |  |
| Rheumatoid arthritis | 6099 (53.2) |
| Other inflammatory arthritis<sup>†</sup> | 1959 (17.1) |
| Giant cell arteritis and/or polymyalgia rheumatica | 388 (3.4) |
| Systemic lupus erythematosus | 1479 (12.9) |
| AAV and other vasculitides<sup>†</sup> | 268 (2.3) |
| Other rheumatic disease<sup>‡</sup> | 571 (5.0) |
| Multiple rheumatic diseases | 704 (6.1) |
| **Immunomodulatory medications<sup>§</sup>** |  |
| Hydroxychloroquine or chloroquine | 3751 (32.7) |
| Methotrexate | 4058 (35.4) |
| Mycophenolate mofetil or mycophenolic acid | 517 (4.5) |
| Other csDMARD<sup>¶</sup> | 1829 (16.9) |
| tsDMARD/JAK inhibitor | 702 (6.1) |
| Anti-CD20 monoclonal antibody | 278 (2.4) |
| TNF inhibitor | 3142 (27.4) |
| IL-6 inhibitor | 458 (4.0) |
| CTLA-4 Ig | 370 (3.2) |
| IL-17, IL-12/23, or IL-23 inhibitor | 597 (5.2) |
| Other bDMARD<sup>#</sup> | 139 (1.2) |
| Cyclophosphamide | 69 (0.6) |
| Oral glucocorticoid | 1162 (10.1) |

AAV, anti-neutrophil cytoplasmic antibody-associated vasculitis; DMARD, disease-modifying antirheumatic drug; bDMARD, biologic DMARD; tsDMARD, targeted synthetic DMARD; JAK, Janus kinase; csDMARD, conventional synthetic DMARD; TNF, tumor necrosis factor; IL, interleukin; CTLA-4 Ig, cytotoxic T-lymphocyte associated protein 4 immunoglobulin
\*Other inflammatory arthritis includes psoriatic arthritis, axial spondyloarthritis, and juvenile idiopathic arthritis
†Other vasculitis includes Takayasu arteritis, Behcet disease, and other miscellaneous vasculitis
‡Other rheumatic disease includes Sjogren's syndrome, systemic sclerosis, mixed connective tissue disease, antiphospholipid antibody syndrome, and idiopathic inflammatory myositis)
§All medications that a patient was receiving were included; thus, numbers do not add up to 100%. The most recent biologic DMARD prescribed prior to the index date was considered to be the active medication.
¶Other csDMARD includes leflunomide, azathioprine, sulfasalazine, apremilast, cyclosporine, and tacrolimus

For their primary vaccination, 6080 (53%) patients received the Pfizer-BioNTech vaccine, 4588 (40%) received Moderna, and 800 (7%) received Johnson & Johnson-Janssen. The majority of people were initially vaccinated when the vaccines first became available, before March 31, 2021 (6195, 54%). COVID-19 infection prior to receipt of the initial SARS-CoV-2 vaccine was observed in 560 (5%) patients.

### Factors associated with COVID-19 breakthrough infection

In our study sample, there were 96,867 person-months of follow-up (median 8.7 [IQR 7.9, 9.5] months per person). During this time, there were 251 breakthrough infections, yielding an incidence rate of 2.6 (2.3, 2.9) per 1,000 person-months. Fourteen patients developed COVID-19 infection between the index date and day 14 following their vaccination. The median time from index date to breakthrough infection was 6.3 months (IQR 3.8, 7.8).

There was no strong association of age, sex, body mass index, smoking status, race, or ethnicity with risk of breakthrough infection. However, there was a statistically significant association of older age with lower risk of breakthrough infection (aHR 0.98 per year, 95% CI: 0.98, 0.99). Those with higher CCI scores had a higher risk of breakthrough infection in both unadjusted and adjusted analyses (aHR 1.10 per unit, 95% CI: 1.07, 1.13). COVID-19 infection prior to the index date was associated with a non-statistically significant trend toward a lower risk of breakthrough infection (aHR 0.71, 95% CI: 0.36, 1.39).

In both unadjusted and multivariable adjusted models, compared to antimalarial monotherapy, multiple medications including anti-CD20 monoclonal antibodies (aHR 5.20, 95% CI: 2.85, 9.48), CTLA-4 Ig (aHR 3.52, 95% CI: 1.90, 6.51), mycophenolate (aHR 2.31, 95% CI: 1.25, 4.27), other csDMARDs (including leflunomide, azathioprine, sulfasalazine, apremilast, cyclosporine, or tacrolimus) (aHR 2.21 95% CI: 1.30, 3.75), IL-6 inhibitors (aHR 2.15, 95% CI: 1.09, 4.24), and TNF inhibitors (aHR 1.70, 95% CI: 1.09, 2.66) were associated with higher risk of breakthrough infection (**Table 3; Figure 2**). JAK inhibitor use was associated with a trend toward higher risk of infection in unadjusted analysis (HR 1.92, 95% CI: 0.96, 3.85) that was statistically significant in the multivariable adjusted model (aHR 2.02, 95% CI: 1.01, 4.06). There was no statistically significant association observed with other medication use, including methotrexate or IL-17, 12/23, or 23 inhibitors, with the risk of breakthrough infection. Those using systemic glucocorticoids had a similar risk of breakthrough infection compared to those not receiving glucocorticoids (aHR 1.00, 95% CI: 0.67, 1.50).

**Table 3.**
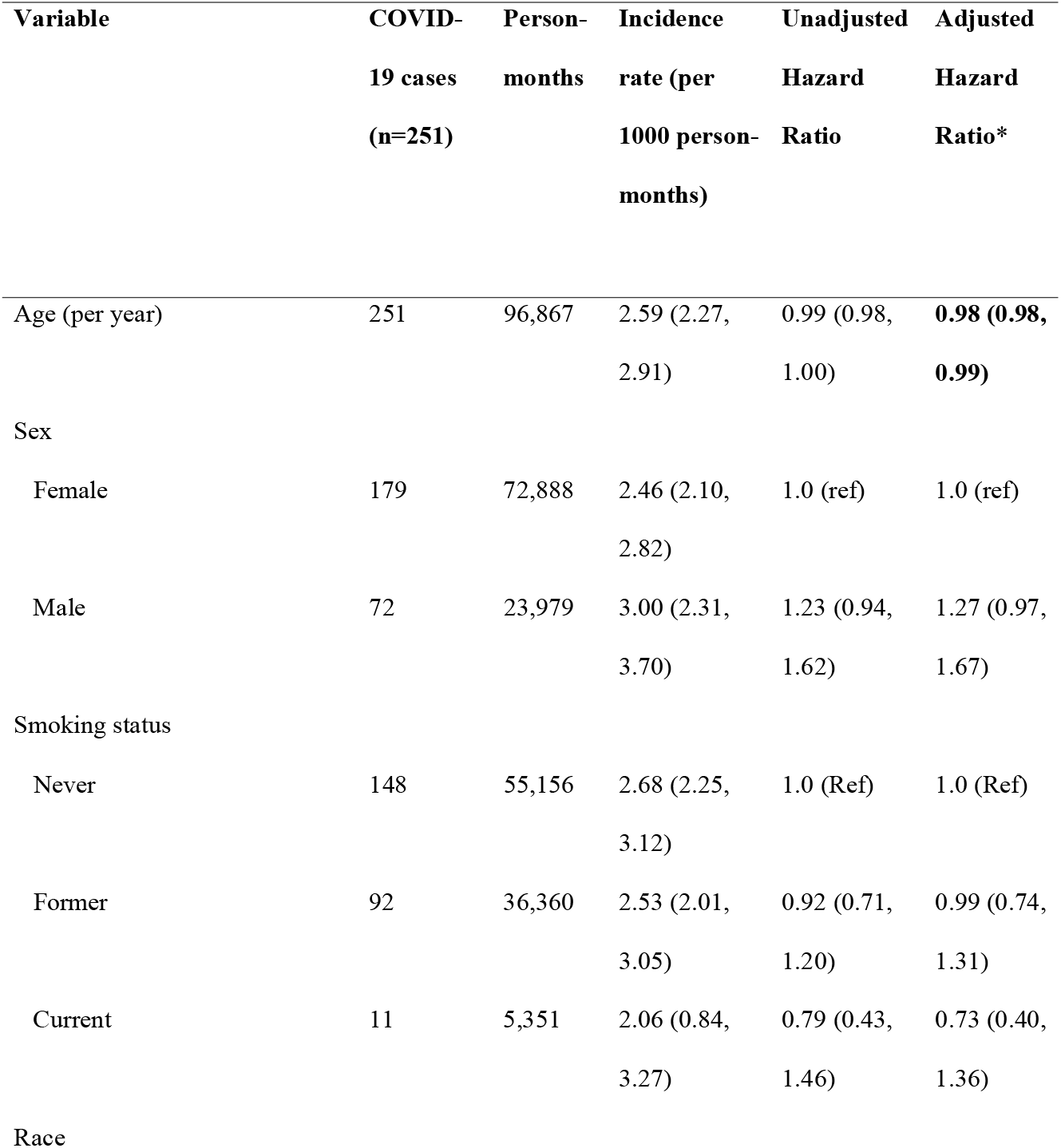
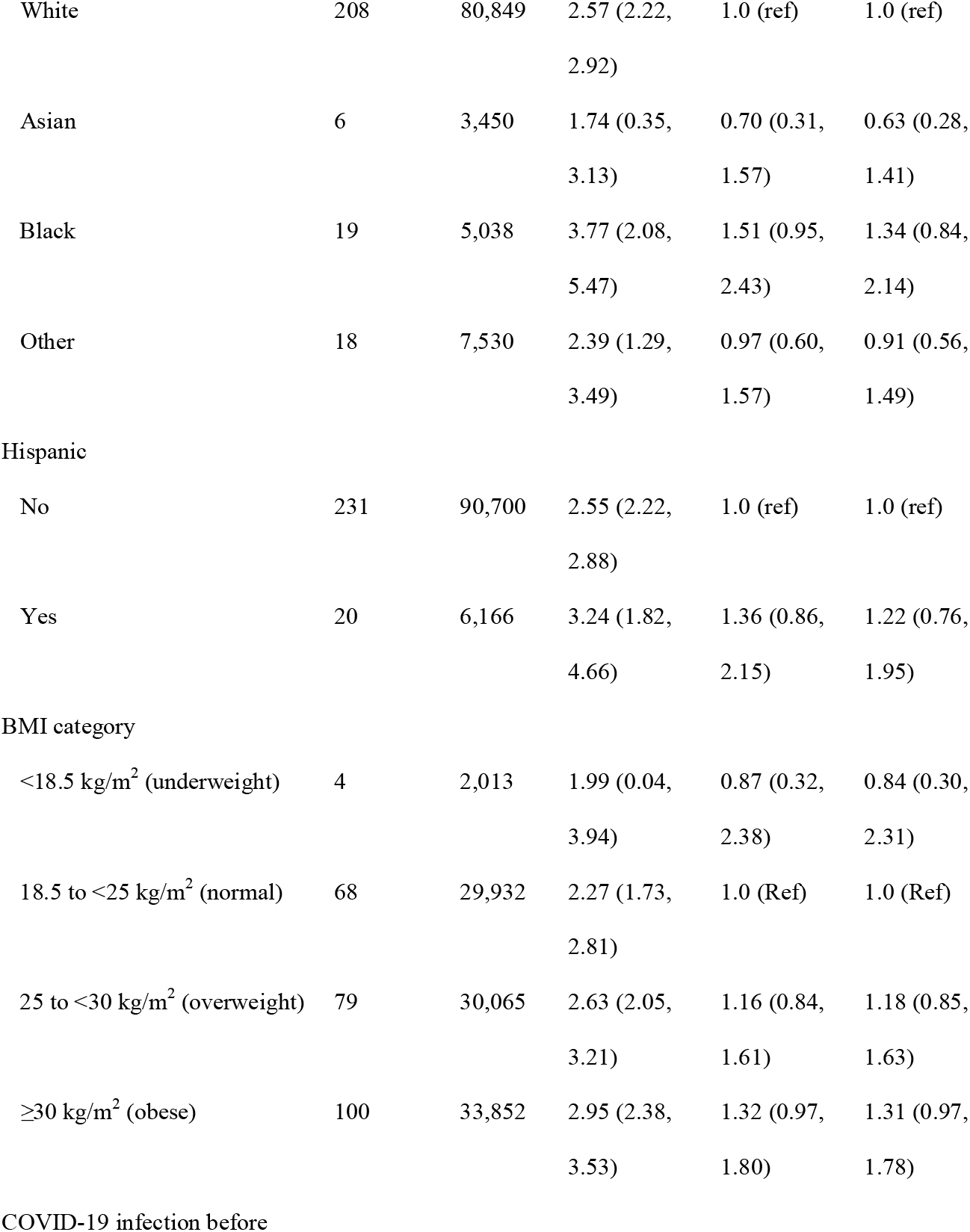
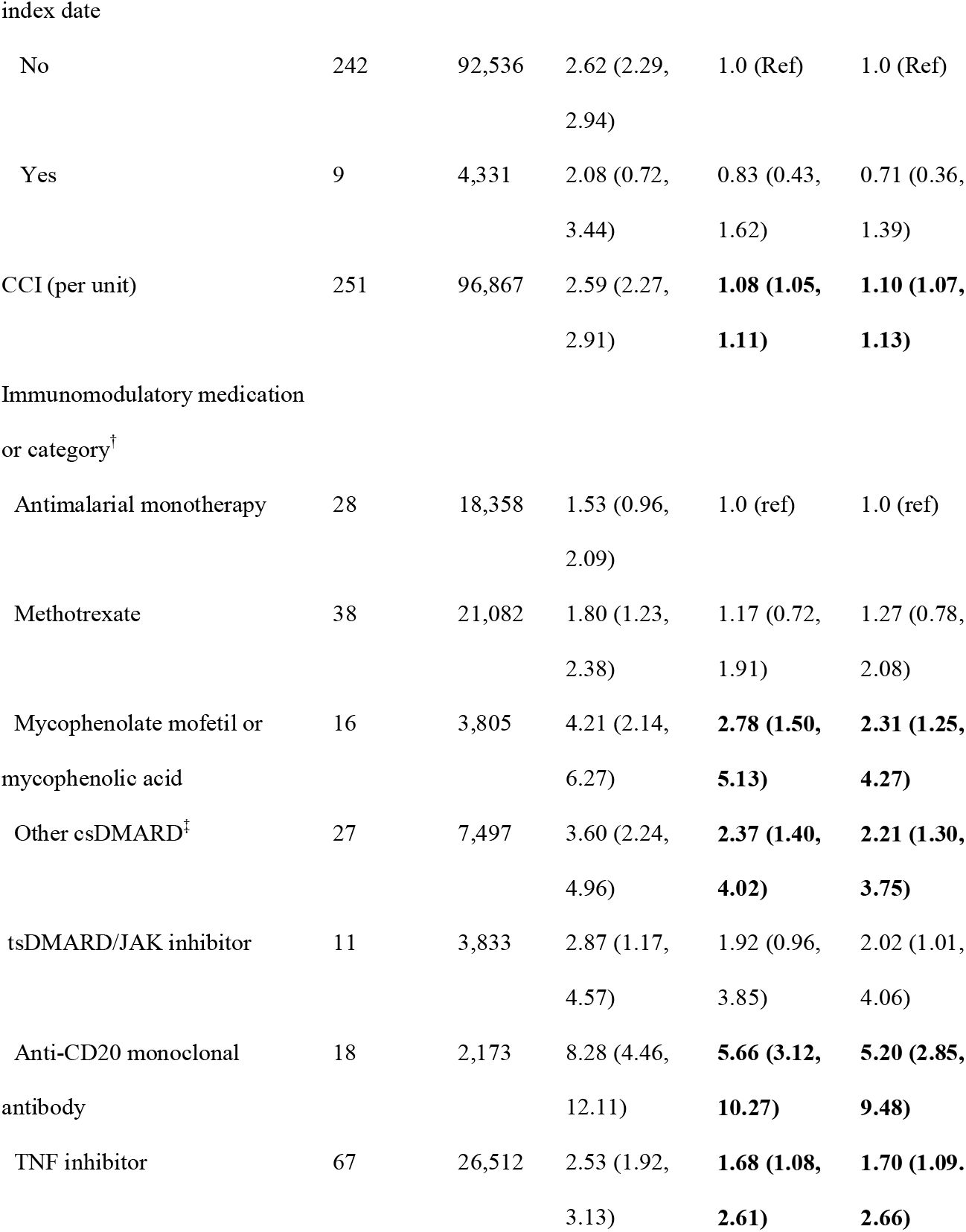
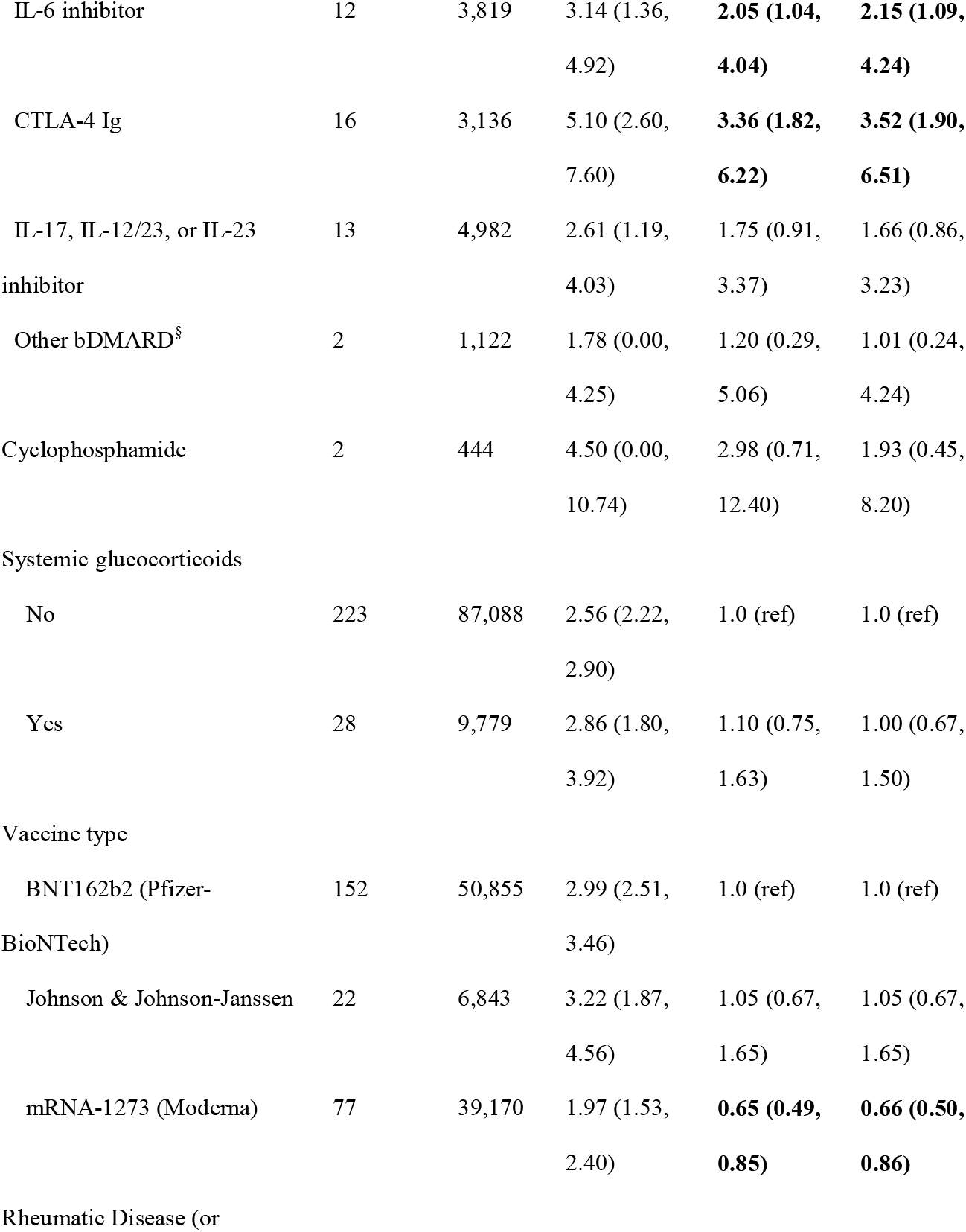
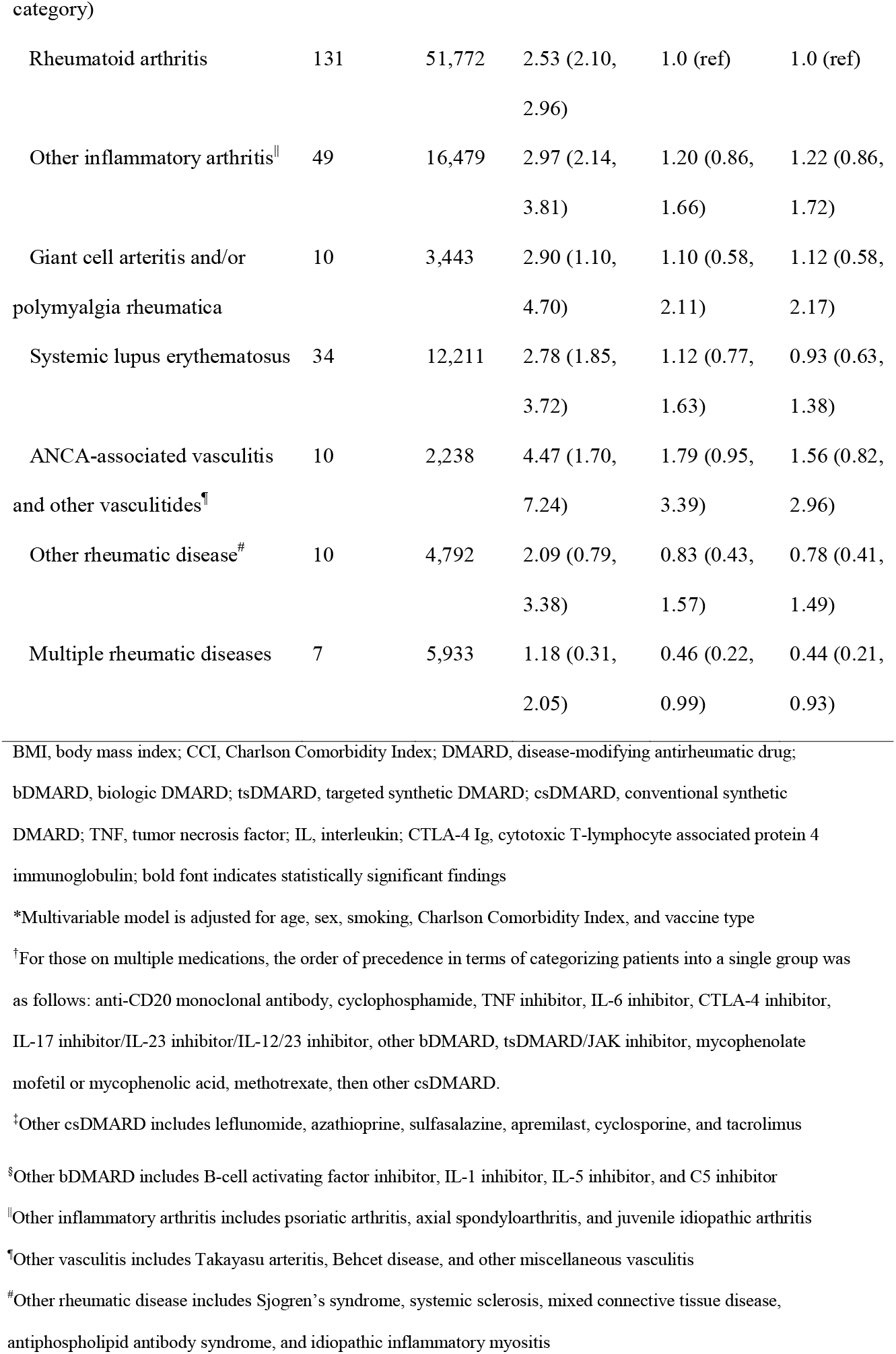
Baseline factors at the time of SARS-CoV-2 vaccination and their associations with COVID-19 breakthrough infection.

**Figure 2.**
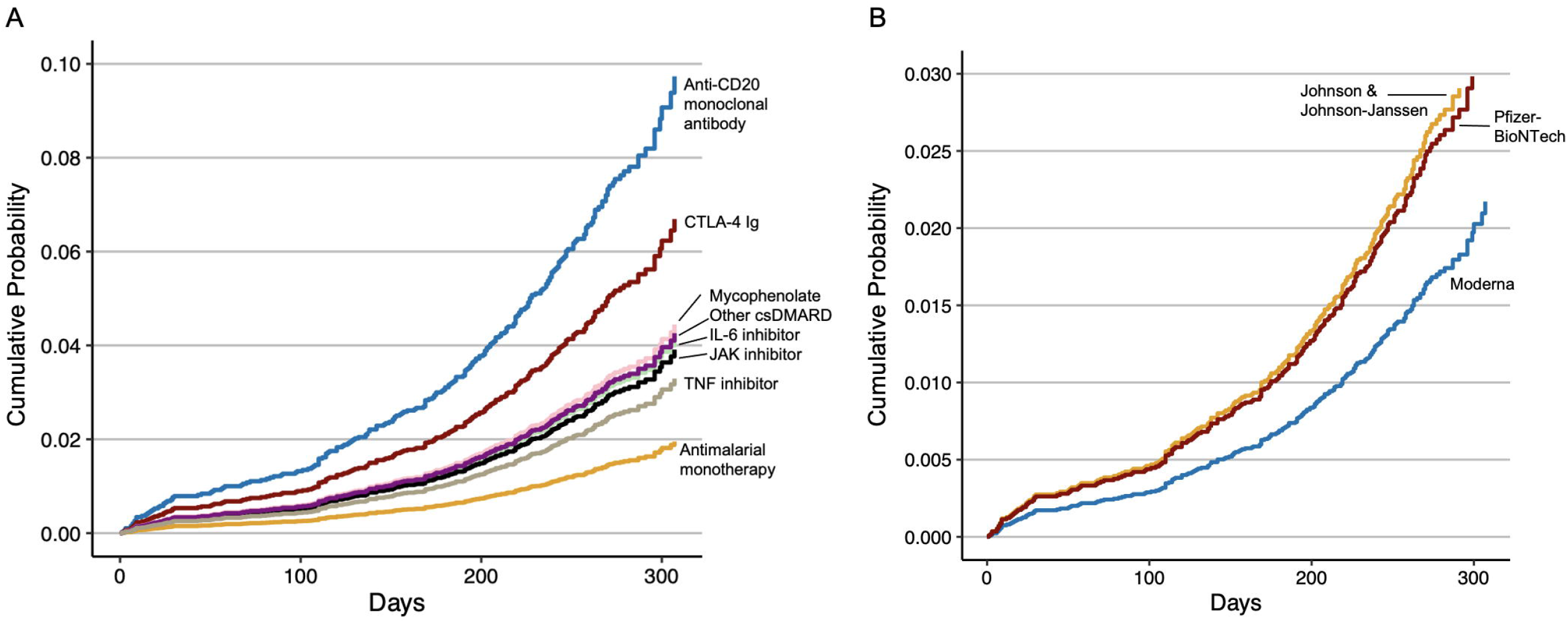
Cumulative probability of COVID-19 breakthrough infection after vaccination, by immunomodulatory medication and vaccine type. CTLA-4 Ig, cytotoxic T-lymphocyte associated protein 4 immunoglobulin; DMARD, disease-modifying antirheumatic drug; IL, interleukin; JAK, Janus kinase; TNF, tumor necrosis factor Cumulative probability of breakthrough infection after the date of initial vaccine series (two doses of either Moderna or Pfizer-BioNTech or one dose of Johnson & Johnson-Janssen) using the multivariable model adjusted for age, sex, smoking, Charlson Comorbidity Index, and vaccine type. A. Cumulative probability in those receiving antimalarial monotherapy, JAK inhibitors, IL-6 inhibitors, CTLA-4 Ig, or anti-CD20 monoclonal antibodies. B. Cumulative probability in those who received the Pfizer-BioNTech, Moderna, or Johnson & Johnson-Janssen vaccine.

Compared to those who received Pfizer-BioNTech, those who received the Moderna vaccine had a lower risk of breakthrough infection (aHR 0.66, 95% CI: 0.50, 0.86). Those who received the Johnson & Johnson-Janssen vaccine had a similar risk of breakthrough infection when compared to those who received Pfizer-BioNTech (aHR 1.05, 95% CI: 0.67, 1.65). There was no association of any particular SARD diagnosis with risk of breakthrough infection.

Of the 251 patients with COVID-19 breakthrough infection, 27 (11%) were hospitalized and 7 (3%) died within 30 days of infection. Low numbers of severe outcomes precluded our ability to evaluate associations of immunomodulatory medications and other factors with severe outcomes.

### Subgroup and sensitivity analyses

When limiting the outcome definition to those with infection at least 14 days after the index date (n=237 breakthrough infections), results were similar, with significantly increased risk of breakthrough infection in those receiving anti-CD20 monoclonal antibodies, CTLA-4 Ig, mycophenolate, other csDMARDs, or TNF inhibitors compared to those receiving antimalarial monotherapy. Those receiving IL-6 inhibitors or JAK inhibitors had a trend toward higher risk, but this did not reach statistical significance (**Supplementary Table 3**). When censoring at the time of any additional vaccine dose (n=194 breakthrough infections), results were overall consistent with the primary analysis, with anti-CD20 monoclonal antibodies (aHR 4.97, 95% CI: 2.36, 10.48) and CTLA-4 Ig (aHR 4.64, 95% CI: 2.35, 9.16) showing the strongest association with increased risk of breakthrough infection (**Supplementary Table 4**).

When extending follow-up through February 22, 2022 to include the initial phase of the Omicron wave in Massachusetts (n=896 breakthrough infections), anti-CD20 monoclonal antibodies (aHR 2.30, 95% CI: 1.60, 3.29), TNF inhibitors (aHR 1.24, 95% CI: 1.01, 1.52), and CTLA-4 Ig (aHR 1.84, 95% CI: 1.31, 2.48) remained associated with a higher risk of breakthrough infection. However, the associations of mycophenolate, other csDMARD, JAK inhibitor, or IL-6 inhibitor use with breakthrough infection were no longer statistically significant (**Supplementary Table 5**). There was no association of specific rheumatic disease diagnosis with risk of breakthrough infection in any of the sensitivity analyses. Receipt of the Moderna vaccine was associated with a lower risk of breakthrough infection compared to the Pfizer-BioNTech vaccine in each sensitivity analysis.

## Discussion

In this multicenter retrospective cohort study of SARD patients using immunomodulatory medications in the pre-Omicron era, we observed a higher risk of COVID-19 breakthrough infection among those using mycophenolate, JAK inhibitors, anti-CD20 monoclonal antibodies, TNF inhibitors, IL-6 inhibitors, CTLA-4 Ig, and other csDMARDs including leflunomide, azathioprine, sulfasalazine, apremilast, cyclosporine, or tacrolimus. Those who received the Moderna vaccine had a lower risk of breakthrough infection. The assembled cohort used in this study is reflective of the spectrum of SARD diagnoses and immunomodulator use one might encounter in a typical rheumatology practice. Our findings suggest that breakthrough infection risk varies substantially depending on immunomodulator use or vaccine type in addition to comorbidity burden. Patients and clinicians should be aware of how these factors may impact infection risk. Despite these observations, the small number of severe outcomes may provide reassurance that vaccinations offer protection against hospitalization and death even among immunosuppressed patients with SARDs.^28^

The risk of breakthrough infection among SARDs has been of high concern since early studies reported the association of immunomodulator use with a blunted antibody response to COVID-19 vaccines.^11^ While some studies have suggested that a retained cellular response may be observed among those with a blunted antibody response, the known importance of antibodies both in preventing infection and in the early response to infection have left many concerned about breakthrough risk in SARD patients.^29,30^

Previous studies have found that patients with immune-mediated diseases, especially those on immunomodulators, are at higher risk of COVID-19 breakthrough infections. Sun *et al* found that patients with rheumatoid arthritis had 20% higher risk of COVID-19 after vaccination using a large US hospital network.^19^ However, Boekel *et al* found no difference in risk for breakthrough infection between patients with immune-mediated inflammatory diseases and controls in the Netherlands.^18^ Importantly, the subset on anti-CD20 monoclonal antibodies had a higher risk of breakthrough infection in that study and an increased risk of breakthrough infection as well as severe outcomes in others.^17,18,31,32^ These findings seemingly conflict with the observed association of multiple DMARDs, not just B cell-depleting agents, with an impaired humoral response to vaccines; indeed, a higher risk of breakthrough infection is associated with a blunted antibody response.^10-14,18^ Our study expands on previous studies of breakthrough infection risk by leveraging a large cohorts of SARD patients using a variety of immunomodulators and vaccinated against COVID-19, most often with an mRNA vaccine.

Our findings suggest that the impaired humoral response observed with multiple classes of immunomodulators, not just B cell depletion, likely predisposed these patients to a higher risk of breakthrough infection. Indeed, previous studies have found that many of the immunomodulators we found to be associated with a higher risk of breakthrough infection, including methotrexate, mycophenolate, and TNF-inhibitors, have been associated with a blunted antibody response to either the COVID-19 vaccine or other types of vaccines.^10-14,33^ Importantly, studies of the immune response to COVID-19 vaccines have often been limited by a small number of patients on less frequently used immunomodulators, such as abatacept and JAK inhibitors, though there is biological plausibility to suspecting that they blunt the immune response.^34^ Our results highlight the importance of understanding the immunologic response to vaccination in patients using immunomodulators reflecting the spectrum of use in day-to-day clinical practice to inform risk mitigation strategies.

We found no association with any specific SARD diagnoses and risk of breakthrough infection. This is consistent with our hypothesis that the main contributors to breakthrough infection risk are the use of certain immunosuppressive medications, as well as comorbidity burden and sociodemographic features.

We also found that patients who received the Moderna vaccine had lower risk of breakthrough infection compared to those who received the Pfizer-BioNTech vaccine. Data have supported a potentially greater effectiveness of the Moderna vaccine for preventing COVID-19 infection compared to the Pfizer-BioNTech vaccine. A study from the Centers for Disease Control and Prevention found that the Moderna vaccine is 86% effective for preventing COVID-19-associated hospitalizations compared with 75% for the Pfizer-BioNTech vaccine, perhaps related to the higher antibody levels associated with Moderna vaccination.^35,36^ Though we adjusted for a number of important potential confounders, an important limitation of our analysis is that the Pfizer-BioNTech vaccine received emergency authorization for use in the US one week before the Moderna vaccine so Pfizer-BioNTech recipients may have experienced vaccine waning sooner, contributing to the observed higher risk. However, similar observations were made in a study by Hernan *et al* which rigorously accounted for these differences.^35^ Additional studies in the SARD population are needed to determine the potential benefits associated with one vaccine type over another to guide future vaccine strategies in this vulnerable population. The most important way to prevent breakthrough infection is to get vaccinated with an effective vaccine. There were too few subjects who received Johnson & Johnson-Janssen to generate robust results and this vaccine is no longer recommended in the US.

Our study has multiple strengths. First, we systematically identified patients with SARDs prescribed immunomodulatory medications prior to vaccination using a rigorous algorithm (90% PPV) applied in a large healthcare system’s data warehouse. Second, we identified test-confirmed COVID-19 breakthrough infection using data from both PCR tests as well as antigen tests administered both in the healthcare setting and at home. Third, details regarding comorbidities and medications prescribed were available. Fourth, in contrast to previous studies, over 90% of patients in our cohort received an mRNA-based vaccine, with a similar proportion receiving Pfizer-BioNTech and Moderna.

Despite these strengths, our study has certain limitations. First, our outcome of COVID-19 breakthrough infection was obtained from testing data (either PCR or antigen) from the EHR and thus patients who were either tested outside of our healthcare system (and outside of other affiliated sites from which our EHR pulls data) or those who performed self-testing at home and did not report their test result may not be captured. However, we do not suspect that testing was differentially distributed across our exposures of interest given the wide availability of testing in this geographic region during our study period. Indeed, the vast majority of diagnoses were diagnosed and managed as outpatients. Second, rheumatic disease diagnosis was ascertained using an algorithm involving multiple ICD-10 codes as well as immunomodulator use. While we did not manually review all charts to confirm the rheumatic disease diagnosis, other studies have shown that these methods have good specificity for identifying patients with rheumatic diseases, as confirmed by our random review of a subset of cases included in this study.^23^ Third, we did not have data about rheumatic disease activity, medication interruption around the time of vaccination, and spike protein antibody response, all of which may have impacted results. Our findings identify important avenues for prioritizing future investigations regarding vaccine efficacy in this population. Fourth, multivariable adjustment was limited in some subgroup and sensitivity analyses by small sample sizes. Fifth, our observed rates of breakthrough infections may be an underestimate, due to asymptomatic or mild cases for which people may not seek testing. However, we limited our study population to Massachusetts residents who would be most likely to seek testing within MGB or other healthcare systems with linked data systems to minimize this potential bias. Sixth, we used antimalarial monotherapy as the reference group when assessing the risk of breakthrough infection with different DMARDs which may have biased our observations towards higher risk given potential confounding by indication and lack of immunosuppression from antimalarials. However, we adjusted for multiple covariates and selected an active comparator, rather than non-use, that we do not expect to affect vaccine response, whereas selecting an alternative comparator such as methotrexate, which may affect vaccine response, would have made comparisons difficult to interpret.

In summary, we found associations between certain DMARDs with a higher risk of breakthrough infection. Additionally, the Moderna vaccine may provide better protection than other vaccines in patients with rheumatic diseases. These findings provide important avenues for future investigation and will be clinically useful for identifying patients in whom additional mitigation or preventative strategies may be most important.

## Data Availability

Deidentified data produced in the present study are available upon reasonable request to the authors

## Notes

**<u>Financial Support</u>** NJP is supported by the Rheumatology Research Foundation. JAS is funded by NIH/NIAMS (grant numbers, R01 AR077607, P30 AR070253, and P30 AR072577), the R. Bruce and Joan M. Mickey Research Scholar Fund, and the Llura Gund Award for Rheumatoid Arthritis Research and Care. ZSW is funded by NIH/NIAMS [K23AR073334 and R03AR078938].

**<u>Declaration of interests</u>** NJP reports consulting fees from FVC Health unrelated to the current work. JAS reports research support from Bristol Myers Squibb and consultancy fees from AbbVie, Amgen, Boehringer Ingelheim, Bristol Myers Squibb, Gilead, Inova Diagnostics, Janssen, Optum, and Pfizer. ZSW reports research support from Bristol-Myers Squibb and Principia/Sanofi and consulting fees from Zenas Biopharma, Horizon, Sanofi, Shionogi, Viela Bio, and MedPace. All other authors report no competing interests.

### Competing Interest Statement

NJP reports consulting fees from FVC Health unrelated to the current work. JAS reports research support from Bristol Myers Squibb and consultancy fees from AbbVie, Amgen, Boehringer Ingelheim, Bristol Myers Squibb, Gilead, Inova Diagnostics, Janssen, Optum, and Pfizer. ZSW reports research support from Bristol-Myers Squibb and Principia/Sanofi and consulting fees from Zenas Biopharma, Horizon, Sanofi, Shionogi, Viela Bio, and MedPace. All other authors report no competing interests.

### Funding Statement

NJP is supported by the Rheumatology Research Foundation. JAS is funded by NIH/NIAMS (grant numbers, R01 AR077607, P30 AR070253, and P30 AR072577), the R. Bruce and Joan M. Mickey Research Scholar Fund, and the Llura Gund Award for Rheumatoid Arthritis Research and Care. ZSW is funded by NIH/NIAMS [K23AR073334 and R03AR078938].

### Author Declarations

Institutional Review Board of Mass General Brigham gave ethical approval for this work. (2020P000833)

