## Supplemental File for "Factors Associated with COVID-19 Breakthrough Infection in the Pre-Omicron Era Among Vaccinated Patients with Rheumatic Diseases: A Cohort Study"

**Supplementary Table 1**. Rheumatic disease terms used to identify patients with systemic rheumatic disease and their associated ICD-10 codes

| **Category** | **Rheumatic Disease (ICD-10 codes)** |
| --- | --- |
| Inflammatory arthritis | - Rheumatoid arthritis (M05%, M06%) - Inflammatory arthritis or inflammatory polyarthropathy (M06.4) - Juvenile idiopathic arthritis (M08.20) - Psoriatic arthritis or arthropathic psoriasis (L40.50) - Ankylosing spondylitis (M45.9) |
| Vasculitis | - Anti-neutrophil cytoplasmic antibody-associated vasculitis: granulomatosis with polyangiitis, eosinophilic granulomatosis with polyangiitis, microscopic polyangiitis (M31.3, M31.7, M30.0) - Kawasaki disease (M30.3) - Takayasu arteritis (M31.4) - Polyarteritis nodosa (M30.0) - Giant cell arteritis (M31.6) - Polymyalgia rheumatica (M35.3) - Behçet disease (M35.2) - Unspecified arteritis (I77.6) |
| Other Systemic Autoimmune Diseases | - Systemic lupus erythematosus (M32%) - Sjogren’s syndrome (M35.0) - Idiopathic inflammatory myositis: dermatomyositis, polymyositis, statin-associated autoimmune myositis, unspecified myositis (G72.49, G72.41, M33) - Systemic sclerosis (M34.0, M34.1, M34.8%, M34.9) - Mixed connective tissue disease (M35.1) - Antiphospholipid syndrome (D68.61) |

ICD = international classification of disease, 10^th^ revision codes. % indicates a wild character to capture all instances of a given code.

**Supplementary Table 2**. Immunomodulatory medications used to identify patients with systemic rheumatic disease

| **Category and generic name** | **Brand name(s)** | **Mechanism of action (for targeted therapy)** | **Route** | **CPT code if intravenous** |
| --- | --- | --- | --- | --- |
| **Glucocorticoids** |  |  |  |  |
| Prednisone (minimum of 30 pills) | Deltasone  Prednicot  Prednisone Intensol  Rayos  Sterapred  Sterapred DS |  | Oral |  |
| Methylprednisolone (minimum of 30 pills) | Medrol |  | Oral |  |
| **Conventional Synthetic DMARDs** |  |  |  |  |
| Azathioprine | Imuran  Azasan |  | Oral |  |
| Methotrexate | Otrexup  Rasuvo  Rheumatrex  Trexall |  | Oral or subQ |  |
| Leflunomide | Arava |  | Oral |  |
| Mycophenolic acid | CellCept |  | Oral |  |
| Mycophenolate mofetil | Myfortic |  | Oral |  |
| Sulfasalazine | Azulfidine |  | Oral |  |
| Hydroxychloroquine | Plaquenil |  | Oral |  |
| Chloroquine | Aralen |  | Oral |  |
| **Targeted Synthetic DMARDs** |  |  |  |  |
| Tofacitinib | Xeljanz | JAK inhibitor | Oral |  |
| Baricitinib | Olumiant | JAK inhibitor | Oral |  |
| Upadacitinib | Rinvoq | JAK inhibitor | Oral |  |
| **Biologic DMARDs** |  |  |  |  |
| Rituximab | Rituxan  Truxima  Ruxience | Anti-CD20 monoclonal antibody | IV | J9310  Q5115  Q5119 |
| Ocrelizumab | Ocrevus | Anti-CD20 monoclonal antibody | IV | J2350 |
| Abatacept | Orencia | CTLA-4 Ig | subQ or IV | J0129 |
| Infliximab | Remicade  Inflectra  Renflexis  Avsola | TNF inhibitor | IV | J1745  Q5103 Q5104  Q5121 |
| Etanercept | Enbrel | TNF inhibitor | subQ |  |
| Adalimumab | Humira | TNF inhibitor | subQ |  |
| Certolizumab | Cimzia | TNF inhibitor | subQ or IV | J0717 |
| Golimumab | Simponi | TNF inhibitor | subQ or IV | J1602 |
| Anakinra | Kineret | IL-1 inhibitor | subQ or IV | n/a (coded under “other biologic” J3490 or J3590 but doesn’t seem to have its own) |
| Canakinumab | Ilaris | IL-1 inhibitor | subQ or IV | J0638 |
| Mepolizumab | Nucala | IL-5 inhibitor | subQ or IV | J2182 |
| Benralizumab | Fasenra | IL-5 inhibitor | subQ or IV | J0517 |
| Tocilizumab | Actemra | IL-6 inhibitor | subQ or IV | J3262 |
| Sarilumab | Kevzara | IL-6 inhibitor | subQ |  |
| Secukinumab | Cosentyx | IL-17A inhibitor | subQ |  |
| Ixekizumab | Taltz | IL-17A inhibitor | subQ |  |
| Ustekinumab | Stelara | IL-12/23 inhibitor | subQ or IV | J3358 |
| Guselkumab | Tremfya | IL-23 inhibitor | subQ |  |
| Belimumab | Benlysta | BLyS inhibitor | subQ or IV | J0490 |
| Eculizumab | Soliris | C5 inhibitor | IV | J1300 |
| **Other** |  |  |  |  |
| Tacrolimus | Prograf  Envarsus  Astagraf  Hecoria |  | Oral or IV | J7525 |
| Cyclosporine | Gengraf  Neoral  Sandimmune |  | Oral or IV | J7516 |
| Apremilast | Otezla | PDE4 inhibitor | Oral |  |
| Cyclophosphamide | Cytoxan |  | Oral or IV | J9070 |

CPT, Current Procedural Terminology; CTLA-4, cytotoxic T-lymphocyte-associated protein 4; DMARD, disease-modifying anti-rheumatic drug; IL, interleukin; JAK, Janus kinase; PDE, phosphodiesterase; TNF, tumor necrosis factor

**Supplementary Table 3**. Baseline factors at the time of SARS-CoV-2 vaccination and their associations with breakthrough infection, limiting outcome definition to those with infection at least 14 days after the index date.

| **Variable** | **COVID-19 cases (n=237)** | **Person-months** | **Incidence rate (per 1000 person-months)** | **Unadjusted Hazard Ratio** | **Multivariable Hazard Ratio*** |
| --- | --- | --- | --- | --- | --- |
| Age (per year) | 237 | 96,986 | 2.44 (2.13, 2.75) | 0.99 (0.98, 1.00) | **0.98 (0.98, 0.99)** |
| Sex |  |  |  |  |  |
| Female | 168 | 72989 | 2.30 (1.95, 2.65) | 1.0 (ref) | 1.0 (ref) |
| Male | 69 | 23996 | 2.88 (2.20, 3.55) | 1.26 (0.95, 1.66) | 1.30 (0.98, 1.72) |
| Smoking status |  |  |  |  |  |
| Never | 140 | 55,220 | 2.54 (2.12, 2.96) | 1.0 (ref) | 1.0 (ref) |
| Former | 86 | 36,414 | 2.36 (1.86, 2.86) | 0.91 (0.70, 1.19) | 0.98 (0.73, 1.31) |
| Current | 11 | 5,351 | 2.06 (0.84, 3.27) | 0.84 (0.46, 1.56) | 0.78 (0.42, 1.45) |
| Race |  |  |  |  |  |
| White | 196 | 80957 | 2.42 (2.08, 2.76) | 1.0 (ref) | 1.0 (ref) |
| Asian | 6 | 3450 | 1.74 (0.35, 3.13) | 0.75 (0.33, 1.68) | 0.67 (0.29, 1.51) |
| Black | 18 | 5039 | 3.57 (1.92, 5.22) | 1.53 (0.94, 2.48) | 1.36 (0.84, 2.20) |
| Other | 17 | 7540 | 2.25 (1.18, 3.33) | 0.98 (0.60, 1.60) | 0.92 (0.56, 1.52) |
| Hispanic |  |  |  |  |  |
| No | 218 | 90809 | 2.40 (2.08, 2.72) | 1.0 (ref) | 1.0 (ref) |
| Yes | 19 | 6177 | 3.08 (1.69, 4.46) | 1.38 (0.87, 2.21) | 1.23 (0.76, 2.00) |
| BMI category |  |  |  |  |  |
| <18.5 kg/m^2^ (underweight) | 4 | 2,013 | 1.99 (0.04, 3.94) | 0.90 (0.33, 2.46) | 0.87 (0.32, 2.41) |
| 18.5 to <25 kg/m^2^ (normal) | 66 | 29,952 | 2.20 (1.67, 2.74) | 1.0 (ref) | 1.0 (ref) |
| 25 to <30 kg/m^2^ (overweight) | 75 | 30,103 | 2.49 (1.93, 3.06) | 1.14 (0.82, 1.59) | 1.15 (0.83, 1.59) |
| ≥30 kg/m^2^ (obese) | 92 | 33,913 | 2.71 (2.16, 3.27) | 1.25 (0.91, 1.72) | 1.24 (0.91, 1.70) |
| COVID-19 infection before index date |  |  |  |  |  |
| No | 231 | 92,629 | 2.49 (2.17, 2.82) | 1.0 (ref) | 1.0 (ref) |
| Yes | 6 | 4,357 | 1.38 (0.28, 2.48) | 0.58 (0.26, 1.31) | 0.50 (0.22, 1.13) |
| CCI (per unit) | 237 | 96985 | 2.44 (2.13, 2.75) | **1.08 (1.05, 1.11)** | **1.10 (1.06, 1.13)** |
| Immunomodulatory medication or category^†^ |  |  |  |  |  |
| Antimalarial monotherapy | 28 | 18358 | 1.53 (0.96, 2.09) | 1.0 (ref) | 1.0 (ref) |
| Methotrexate | 37 | 21090 | 1.75 (1.19, 2.32) | 1.14 (0.70, 1.86) | 1.24 (0.76, 2.03) |
| Mycophenolate mofetil or mycophenolic acid | 14 | 3820 | 3.67 (1.75, 5.58) | **2.43 (1.28, 4.61)** | **2.01 (1.06, 3.82)** |
| Other csDMARD^‡^ | 25 | 7507 | 3.33 (2.02, 4.64) | **2.19 (1.28, 3.75)** | **2.04 (1.19, 3.50)** |
| tsDMARD/JAK inhibitor | 10 | 3842 | 2.60 (0.99, 4.22) | 1.74 (0.85, 3.58) | 1.84 (0.89, 3.77) |
| Anti-CD20 monoclonal antibody | 16 | 2194 | 7.29 (3.72, 10.87) | **5.01 (2.70, 9.29)** | **4.61 (2.47, 8.61)** |
| TNF inhibitor | 65 | 26532 | 2.45 (1.85, 3.05) | **1.63 (1.05, 2.54)** | **1.63 (1.04, 2.56)** |
| IL-6 inhibitor | 10 | 3838 | 2.61 (0.99, 4.22) | 1.70 (0.83, 3.50) | 1.79 (0.87, 3.69) |
| CTLA-4 Ig | 16 | 3136 | 5.10 (2.60, 7.60) | **3.36 (1.82, 6.22)** | **3.53 (1.90, 6.54)** |
| IL-17, IL-12/23, or IL-23 inhibitor | 11 | 4999 | 2.20 (0.90, 3.50) | 1.48 (0.74, 2.96) | 1.39 (0.69, 2.80) |
| Other bDMARD^§^ | 2 | 1122 | 1.78 (0.00, 4.25) | 1.21 (0.29, 5.08) | 1.01 (0.24, 4.24) |
| Cyclophosphamide | 2 | 444 | 4.50 (0.00, 10.74) | 2.99 (0.72, 12.46) | 1.94 (0.46, 8.26) |
| Systemic glucocorticoids |  |  |  |  |  |
| No | 212 | 87190 | 2.43 (2.10, 2.76) | 1.0 (ref) | 1.0 (ref) |
| Yes | 25 | 9796 | 2.55 (1.55, 3.55) | 1.04 (0.68, 1.57) | 0.95 (0.62, 1.45) |
| Vaccine type |  |  |  |  |  |
| BNT162b2 (Pfizer-BioNTech) | 143 | 50926 | 2.81 (2.35, 3.27) | 1.0 (ref) | 1.0 (ref) |
| Johnson & Johnson-Janssen | 21 | 6851 | 3.07 (1.75, 4.38) | 1.07 (0.67, 1.69) | 1.06 (0.67, 1.68) |
| mRNA-1273 (Moderna) | 73 | 39208 | 1.86 (1.43, 2.29) | **0.65 (0.49, 0.86)** | **0.66 (0.50, 0.87)** |
| Rheumatic Disease (or category) |  |  |  |  |  |
| Rheumatoid arthritis | 122 | 51846 | 2.35 (1.94, 2.77) | 1.0 (ref) | 1.0 (ref) |
| Other inflammatory arthritis^\|\|^ | 46 | 16506 | 2.79 (1.98, 3.59) | 1.21 (0.86, 1.70) | 1.21 (0.84, 1.73) |
| Giant cell arteritis and/or polymyalgia rheumatica | 10 | 3443 | 2.90 (1.10, 4.70) | 1.18 (0.62, 2.26) | 1.21 (0.63, 2.34) |
| Systemic lupus erythematosus | 32 | 12229 | 2.62 (1.71, 3.52) | 1.13 (0.77, 1.67) | 0.94 (0.63, 1.41) |
| ANCA-associated vasculitis and other vasculitides^¶^ | 10 | 2238 | 4.47 (1.70, 7.24) | **1.93 (1.02, 3.66)** | 1.67 (0.88, 3.18) |
| Other rheumatic disease^#^ | 10 | 4792 | 2.09 (0.79, 3.38) | 0.89 (0.47, 1.69) | 0.84 (0.44, 1.60) |
| Multiple rheumatic diseases | 7 | 5933 | 1.18 (0.31, 2.05) | 0.50 (0.23, 1.06) | 0.47 (0.22, 1.01) |

BMI, body mass index; CCI, Charlson Comorbidity Index; DMARD, disease-modifying antirheumatic drug; bDMARD, biologic DMARD; tsDMARD, targeted synthetic DMARD; JAK, Janus kinase; csDMARD, conventional synthetic DMARD; TNF, tumor necrosis factor; IL, interleukin; CTLA-4 Ig, cytotoxic T-lymphocyte associated protein 4 immunoglobulin. Bold font indicates statistically significant results.

*Multivariable model is adjusted for age, sex, smoking, Charlson Comorbidity Index, and vaccine type

^†^For those on multiple medications, the order of precedence in terms of categorizing patients into a single group was as follows: anti-CD20 monoclonal antibody, cyclophosphamide, TNF inhibitor, IL-6 inhibitor, CTLA-4 inhibitor, IL-17 inhibitor/IL-23 inhibitor/IL-12/23 inhibitor, other bDMARD, tsDMARD/JAK inhibitor, mycophenolate mofetil or mycophenolic acid, methotrexate, then other csDMARD.

^‡^Other csDMARD includes leflunomide, azathioprine, sulfasalazine, apremilast, cyclosporine, and tacrolimus

^§^Other bDMARD includes B-cell activating factor inhibitor, IL-1 inhibitor, IL-5 inhibitor, and C5 inhibitor

^||^Other inflammatory arthritis includes psoriatic arthritis, axial spondyloarthritis, and juvenile idiopathic arthritis

^¶^Other vasculitis includes Takayasu arteritis, Behcet disease, and other miscellaneous vasculitis

^#^Other rheumatic disease includes Sjogren’s syndrome, systemic sclerosis, mixed connective tissue disease, antiphospholipid antibody syndrome, and idiopathic inflammatory myositis

**Supplementary Table 4**. Baseline factors at the time of SARS-CoV-2 vaccination and their associations with breakthrough infection, censoring at the time of any additional vaccine dose after baseline (defined as the second of any vaccine for those who received initial Johnson & Johnson-Janssen or third of any vaccine for those who received two of either the Pfizer-BioNTech or Moderna vaccines initially).

| **Variable** | **COVID-19 cases (n=194)** | **Person-months** | **Incidence rate (per 1000 person-months)** | **Unadjusted Hazard Ratio** | **Multivariable Hazard Ratio*** |
| --- | --- | --- | --- | --- | --- |
| Age (per year) | 194 | 81,088 | 2.39 (2.06, 2.73) | 0.99 (0.98, 1.00) | 0.99 (0.98, 1.00) |
| Sex |  |  |  |  |  |
| Female | 140 | 60957 | 2.30 (1.92, 2.68) | 1.0 (ref) | 1.0 (ref) |
| Male | 54 | 20131 | 2.68 (1.97, 3.40) | 1.17 (0.86, 1.60) | 1.20 (0.88, 1.64) |
| Smoking status |  |  |  |  |  |
| Never | 112 | 46,152 | 2.43 (1.98, 2.88) | 1.0 (ref) | 1.0 (ref) |
| Former | 73 | 30,185 | 2.42 (1.86, 2.97) | 0.99 (0.73, 1.32) | 1.02 (0.74, 1.41) |
| Current | 9 | 4,751 | 1.89 (0.66, 3.13) | 0.77 (0.39, 1.53) | 0.72 (0.36, 1.43) |
| Race |  |  |  |  |  |
| White | 157 | 67006 | 2.34 (1.98, 2.71) | 1.0 (ref) | 1.0 (ref) |
| Asian | 5 | 2935 | 1.70 (0.21, 3.20) | 0.74 (0.30, 1.79) | 0.68 (0.28, 1.66) |
| Black | 18 | 4537 | 3.97 (2.13, 5.80) | **1.65 (1.01, 2.70)** | 1.48 (0.91, 2.41) |
| Other | 14 | 6610 | 2.12 (1.01, 3.23) | 0.90 (0.52, 1.56) | 0.85 (0.49, 1.49) |
| Hispanic |  |  |  |  |  |
| No | 177 | 75571 | 2.34 (2.00, 2.69) | 1.0 (ref) | 1.0 (ref) |
| Yes | 17 | 5518 | 3.08 (1.62, 4.55) | 1.34 (0.81, 2.20) | 1.22 (0.73, 2.03) |
| BMI category |  |  |  |  |  |
| <18.5 kg/m^2^ (underweight) | 2 | 1,701 | 1.18 (0.00, 2.81) | 0.60 (0.15, 2.49) | 0.58 (0.14, 2.41) |
| 18.5 to <25 kg/m^2^ (normal) | 47 | 24,744 | 1.90 (1.36, 2.44) | 1.0 (ref) | 1.0 (ref) |
| 25 to <30 kg/m^2^ (overweight) | 65 | 25,034 | 2.60 (1.97, 3.23) | 1.36 (0.94, 1.98) | 1.40 (0.96, 2.02) |
| ≥30 kg/m^2^ (obese) | 80 | 28,678 | 2.79 (2.18, 3.40) | 1.46 (1.02, 2.10) | 1.46 (1.02, 2.09) |
| COVID-19 infection before index date |  |  |  |  |  |
| No | 185 | 77,239 | 2.40 (2.05, 2.74) | 1.0 (ref) | 1.0 (ref) |
| Yes | 9 | 3,849 | 2.34 (0.81, 3.87) | 0.98 (0.50, 1.93) | 0.84 (0.42, 1.66) |
| CCI (per unit) | 194 | 81088 | 2.39 (2.06, 2.73) | **1.08 (1.05, 1.12)** | **1.09 (1.06, 1.13)** |
| Immunomodulatory medication or category^†^ |  |  |  |  |  |
| Antimalarial monotherapy | 21 | 16191 | 1.30 (0.74, 1.85) | 1.0 (ref) | 1.0 (ref) |
| Methotrexate | 30 | 17548 | 1.71 (1.10, 2.32) | 1.37 (0.78, 2.39) | 1.47 (0.84, 2.58) |
| Mycophenolate mofetil or mycophenolic acid | 10 | 3059 | 3.27 (1.24, 5.30) | **2.78 (1.31, 5.91)** | **2.34 (1.11, 4.96)** |
| Other csDMARD^‡^ | 22 | 6364 | 3.46 (2.01, 4.90) | **2.77 (1.52, 5.04)** | **2.64 (1.45, 4.80)** |
| tsDMARD/JAK inhibitor | 9 | 3139 | 2.87 (0.99, 4.74) | **2.40 (1.10, 5.24)** | **2.52 (1.16, 5.51)** |
| Anti-CD20 monoclonal antibody | 11 | 1743 | 6.31 (2.58, 10.04) | **5.49 (2.64, 11.42)** | **4.97 (2.36, 10.48)** |
| TNF inhibitor | 54 | 21757 | 2.48 (1.82, 3.14) | **2.05 (1.24, 3.40)** | **2.09 (1.25, 3.49)** |
| IL-6 inhibitor | 10 | 3111 | 3.21 (1.22, 5.21) | **2.65 (1.25, 5.64)** | **2.75 (1.29, 5.86)** |
| CTLA-4 Ig | 14 | 2550 | 5.49 (2.61, 8.37) | **4.55 (2.31, 8.95)** | **4.64 (2.35, 9.16)** |
| IL-17, IL-12/23, or IL-23 inhibitor | 10 | 4240 | 2.36 (0.90, 3.82) | 1.91 (0.90, 4.06) | 1.88 (0.88, 4.02) |
| Other bDMARD^§^ | 1 | 927 | 1.08 (0.00, 3.19) | 0.91 (0.12, 6.78) | 0.76 (0.10, 5.64) |
| Cyclophosphamide | 1 | 370 | 2.70 (0.00, 8.00) | 2.21 (0.30, 16.35) | 1.45 (0.19, 11.17) |
| Systemic glucocorticoids |  |  |  |  |  |
| No | 174 | 73081 | 2.38 (2.03, 2.73) | 1.0 (ref) | 1.0 (ref) |
| Yes | 20 | 8007 | 2.50 (1.40, 3.59) | 1.06 (0.67, 1.69) | 0.97 (0.60, 1.55) |
| Vaccine type |  |  |  |  |  |
| BNT162b2 (Pfizer-BioNTech) | 114 | 42104 | 2.71 (2.21, 3.20) | 1.0 (ref) | 1.0 (ref) |
| Johnson & Johnson-Janssen | 21 | 6273 | 3.35 (1.92, 4.78) | 1.11 (0.70, 1.77) | 1.11 (0.70, 1.76) |
| mRNA-1273 (Moderna) | 59 | 32712 | 1.80 (1.34, 2.26) | **0.64 (0.47, 0.88)** | **0.66 (0.48, 0.90)** |
| Rheumatic Disease (or category) |  |  |  |  |  |
| Rheumatoid arthritis | 99 | 43024 | 2.30 (1.85, 2.75) | 1.0 (ref) | 1.0 (ref) |
| Other inflammatory arthritis^\|\|^ | 41 | 13890 | 2.95 (2.05, 3.86) | 1.29 (0.90, 1.86) | 1.37 (0.94, 2.01) |
| Giant cell arteritis and/or polymyalgia rheumatica | 7 | 2812 | 2.49 (0.65, 4.33) | 1.07 (0.50, 2.31) | 1.09 (0.50, 2.39) |
| Systemic lupus erythematosus | 27 | 10541 | 2.56 (1.60, 3.53) | 1.10 (0.71, 1.68) | 0.95 (0.61, 1.46) |
| ANCA-associated vasculitis and other vasculitides^¶^ | 6 | 1889 | 3.18 (0.63, 5.72) | 1.36 (0.60, 3.10) | 1.24 (0.54, 2.82) |
| Other rheumatic disease^#^ | 7 | 4030 | 1.74 (0.45, 3.02) | 0.75 (0.35, 1.61) | 0.72 (0.33, 1.55) |
| Multiple rheumatic diseases | 7 | 4903 | 1.43 (0.37, 2.49) | 0.62 (0.29, 1.33) | 0.58 (0.27, 1.24) |

BMI, body mass index; CCI, Charlson Comorbidity Index; DMARD, disease-modifying antirheumatic drug; bDMARD, biologic DMARD; tsDMARD, targeted synthetic DMARD; JAK, Janus kinase; csDMARD, conventional synthetic DMARD; TNF, tumor necrosis factor; IL, interleukin; CTLA-4 Ig, cytotoxic T-lymphocyte associated protein 4 immunoglobulin. Bold font indicates statistically significant results.

*Multivariable model is adjusted for age, sex, smoking, Charlson Comorbidity Index, and vaccine type

^†^For those on multiple medications, the order of precedence in terms of categorizing patients into a single group was as follows: anti-CD20 monoclonal antibody, cyclophosphamide, TNF inhibitor, IL-6 inhibitor, CTLA-4 inhibitor, IL-17 inhibitor/IL-23 inhibitor/IL-12/23 inhibitor, other bDMARD, tsDMARD/JAK inhibitor, mycophenolate mofetil or mycophenolic acid, methotrexate, then other csDMARD.

^‡^Other csDMARD includes leflunomide, azathioprine, sulfasalazine, apremilast, cyclosporine, and tacrolimus

^§^Other bDMARD includes B-cell activating factor inhibitor, IL-1 inhibitor, IL-5 inhibitor, and C5 inhibitor

^||^Other inflammatory arthritis includes psoriatic arthritis, axial spondyloarthritis, and juvenile idiopathic arthritis

^¶^Other vasculitis includes Takayasu arteritis, Behcet disease, and other miscellaneous vasculitis

^#^Other rheumatic disease includes Sjogren’s syndrome, systemic sclerosis, mixed connective tissue disease, antiphospholipid antibody syndrome, and idiopathic inflammatory myositis

**Supplementary Table 5**. Baseline factors at the time of SARS-CoV-2 vaccination and their associations with breakthrough infection, extending the follow-up period through February 22, 2022 to include the initial Omicron wave.

| **Variable** | **COVID-19 cases (n=896)** | **Person-months** | **Incidence rate (per 1000 person-months)** | **Unadjusted Hazard Ratio** | **Multivariable Hazard Ratio*** |
| --- | --- | --- | --- | --- | --- |
| Age (per year) | 896 | 121,477 | 7.38 (6.89, 7.86) | **0.98 (0.98, 0.98)** | **0.98 (0.97, 0.98)** |
| Sex |  |  |  |  |  |
| Female | 704 | 91380 | 7.70 (7.13, 8.27) | 1.0 (ref) | 1.0 (ref) |
| Male | 192 | 30097 | 6.38 (5.48, 7.28) | **0.83 (0.71, 0.98)** | 0.88 (0.75, 1.03) |
| Smoking status |  |  |  |  |  |
| Never | 532 | 69,234 | 7.68 (7.03, 8.34) | 1.0 (ref) | 1.0 (ref) |
| Former | 323 | 45,448 | 7.11 (6.33, 7.88) | 0.89 (0.78, 1.02) | 1.10 (0.94, 1.27) |
| Current | 41 | 6,794 | 6.03 (4.19, 7.88) | 0.82 (0.60, 1.13) | 0.83 (0.60, 1.14) |
| Race |  |  |  |  |  |
| White | 723 | 101225 | 7.14 (6.62, 7.66) | 1.0 (ref) | 1.0 (ref) |
| Asian | 28 | 4355 | 6.43 (4.05, 8.81) | 0.95 (0.65, 1.39) | 0.82 (0.56, 1.20) |
| Black | 61 | 6372 | 9.57 (7.17, 11.98) | **1.43 (1.10, 1.85)** | 1.23 (0.94, 1.60) |
| Other | 84 | 9524 | 8.82 (6.93, 10.71) | **1.33 (1.06, 1.67)** | 1.20 (0.96, 1.51) |
| Hispanic |  |  |  |  |  |
| No | 811 | 113645 | 7.14 (6.65, 7.63) | 1.0 (ref) | 1.0 (ref) |
| Yes | 85 | 7832 | 10.85 (8.55, 13.16) | **1.71 (1.37, 2.15)** | **1.45 (1.15, 1.82)** |
| BMI category |  |  |  |  |  |
| <18.5 kg/m^2^ (underweight) | 15 | 2,520 | 5.95 (2.94, 8.96) | 0.86 (0.51, 1.44) | 0.84 (0.50, 1.41) |
| 18.5 to <25 kg/m^2^ (normal) | 257 | 37,467 | 6.86 (6.02, 7.70) | 1.0 (ref) | 1.0 (ref) |
| 25 to <30 kg/m^2^ (overweight) | 262 | 37,724 | 6.95 (6.10, 7.79) | 1.02 (0.86, 1.21) | 1.08 (0.91, 1.28) |
| ≥30 kg/m^2^ (obese) | 361 | 42,492 | 8.50 (7.62, 9.37) | **1.27 (1.09, 1.50)** | **1.27 (1.08, 1.49)** |
| COVID-19 infection before index date |  |  |  |  |  |
| No | 852 | 115,943 | 7.35 (6.86, 7.84) | 1.0 (ref) | 1.0 (ref) |
| Yes | 44 | 5,534 | 7.95 (5.60, 10.30) | 1.17 (0.87, 1.59) | 1.03 (0.76, 1.40) |
| CCI (per unit) | 896 | 121477 | 7.38 (6.89, 7.86) | **1.05 (1.03, 1.07)** | **1.08 (1.06, 1.09)** |
| Immunomodulatory medication or category^†^ |  |  |  |  |  |
| Antimalarial monotherapy | 151 | 23048 | 6.55 (5.51, 7.60) | 1.0 (ref) | 1.0 (ref) |
| Methotrexate | 161 | 26375 | 6.10 (5.16, 7.05) | 0.91 (0.73, 1.14) | 1.09 (0.87, 1.36) |
| Mycophenolate mofetil or mycophenolic acid | 45 | 4783 | 9.41 (6.66, 12.16) | **1.46 (1.05, 2.04)** | 1.26 (0.90, 1.76) |
| Other csDMARD^‡^ | 74 | 9384 | 7.89 (6.09, 9.68) | 1.21 (0.91, 1.60) | 1.23 (0.93, 1.62) |
| tsDMARD/JAK inhibitor | 26 | 4829 | 5.38 (3.31, 7.45) | 0.84 (0.55, 1.28) | 0.90 (0.59, 1.37) |
| Anti-CD20 monoclonal antibody | 40 | 2723 | 14.69 (10.14, 19.24) | **2.40 (1.69, 3.42)** | **2.30 (1.60, 3.29)** |
| TNF inhibitor | 253 | 33276 | 7.60 (6.67, 8.54) | 1.18 (0.97, 1.45) | **1.24 (1.01, 1.52)** |
| IL-6 inhibitor | 33 | 4791 | 6.89 (4.54, 9.24) | 1.04 (0.71, 1.51) | 1.18 (0.81, 1.73) |
| CTLA-4 Ig | 43 | 3898 | 11.03 (7.74, 14.33) | **1.70 (1.21, 2.39)** | **1.84 (1.31, 2.58)** |
| IL-17, IL-12/23, or IL-23 inhibitor | 43 | 6279 | 6.85 (4.80, 8.90) | 1.08 (0.77, 1.52) | 1.07 (0.76, 1.51) |
| Other bDMARD^§^ | 16 | 1408 | 11.36 (5.80, 16.93) | **1.85 (1.10, 3.11)** | 1.48 (0.88, 2.50) |
| Cyclophosphamide | 9 | 554 | 16.25 (5.63, 26.86) | **2.57 (1.30, 5.06)** | 1.97 (0.98, 3.94) |
| Systemic glucocorticoids |  |  |  |  |  |
| No | 768 | 109292 | 7.03 (6.53, 7.52) | 1.0 (ref) | 1.0 (ref) |
| Yes | 128 | 12184 | 10.51 (8.69, 12.33) | **1.47 (1.22, 1.78)** | **1.47 (1.22, 1.79)** |
| Vaccine type |  |  |  |  |  |
| BNT162b2 (Pfizer-BioNTech) | 490 | 63867 | 7.67 (6.99, 8.35) | 1.0 (ref) | 1.0 (ref) |
| Johnson & Johnson-Janssen | 70 | 8550 | 8.19 (6.27, 10.10) | 1.03 (0.80, 1.32) | 1.05 (0.81, 1.35) |
| mRNA-1273 (Moderna) | 336 | 49060 | 6.85 (6.12, 7.58) | 0.87 (0.76, 1.00) | 0.88 (0.76, 1.01) |
| Rheumatic Disease (or category) |  |  |  |  |  |
| Rheumatoid arthritis | 471 | 64855 | 7.26 (6.61, 7.92) | 1.0 (ref) | 1.0 (ref) |
| Other inflammatory arthritis^\|\|^ | 131 | 20744 | 6.31 (5.23, 7.40) | 0.90 (0.74, 1.09) | 0.88 (0.72, 1.08) |
| Giant cell arteritis and/or polymyalgia rheumatica | 27 | 4281 | 6.31 (3.93, 8.69) | 0.81 (0.55, 1.19) | 0.99 (0.67, 1.46) |
| Systemic lupus erythematosus | 132 | 15366 | 8.59 (7.12, 10.06) | **1.22 (1.01, 1.48)** | 0.91 (0.74, 1.11) |
| ANCA-associated vasculitis and other vasculitides^¶^ | 29 | 2800 | 10.36 (6.59, 14.13) | **1.47 (1.01, 2.14)** | 1.22 (0.84, 1.77) |
| Other rheumatic disease^#^ | 58 | 5995 | 9.67 (7.18, 12.16) | **1.35 (1.03, 1.77)** | 1.21 (0.93, 1.59) |
| Multiple rheumatic diseases | 48 | 7435 | 6.46 (4.63, 8.28) | 0.88 (0.65, 1.18) | 0.88 (0.65, 1.18) |

BMI, body mass index; CCI, Charlson Comorbidity Index; DMARD, disease-modifying antirheumatic drug; bDMARD, biologic DMARD; tsDMARD, targeted synthetic DMARD; JAK, Janus kinase; csDMARD, conventional synthetic DMARD; TNF, tumor necrosis factor; IL, interleukin; CTLA-4 Ig, cytotoxic T-lymphocyte associated protein 4 immunoglobulin. Bold font indicates statistically significant results.

*Multivariable model is adjusted for age, sex, smoking, Charlson Comorbidity Index, and vaccine type

^†^For those on multiple medications, the order of precedence in terms of categorizing patients into a single group was as follows: anti-CD20 monoclonal antibody, cyclophosphamide, TNF inhibitor, IL-6 inhibitor, CTLA-4 inhibitor, IL-17 inhibitor/IL-23 inhibitor/IL-12/23 inhibitor, other bDMARD, tsDMARD/JAK inhibitor, mycophenolate mofetil or mycophenolic acid, methotrexate, then other csDMARD.

^‡^Other csDMARD includes leflunomide, azathioprine, sulfasalazine, apremilast, cyclosporine, and tacrolimus

^§^Other bDMARD includes B-cell activating factor inhibitor, IL-1 inhibitor, IL-5 inhibitor, and C5 inhibitor

^||^Other inflammatory arthritis includes psoriatic arthritis, axial spondyloarthritis, and juvenile idiopathic arthritis

^¶^Other vasculitis includes Takayasu arteritis, Behcet disease, and other miscellaneous vasculitis

^#^Other rheumatic disease includes Sjogren’s syndrome, systemic sclerosis, mixed connective tissue disease, antiphospholipid antibody syndrome, and idiopathic inflammatory myositis
